# Five approaches to the suppression of SARS-CoV-2 without intensive social distancing

**DOI:** 10.1101/2020.07.30.20165159

**Authors:** John M. Drake, Pej Rohani, Kyle Dahlin, Andreas Handel

## Abstract

Initial efforts to mitigate transmission of SARS-CoV-2 relied on intensive social distancing measures such as school and workplace closures, shelter-in-place orders, and prohibitions on the gathering of people. Other non-pharmaceutical interventions for suppressing transmission include active case finding, contact tracing, quarantine, immunity or health certification, and a wide range of personal protective measures. Here we investigate the potential effectiveness of these alternative approaches to suppression. We introduce a conceptual framework represented by two mathematical models that differ in strategy. We find both strategies may be effective, although both require extensive testing and work within a relatively narrow range of conditions. Generalized protective measures such as wearing face masks, improved hygiene, and local reductions in density are found to significantly increase the effectiveness of targeted interventions.

## Introduction

Efforts to control the 2020 COVID-19 pandemic have resulted in unprecedented economic impacts. The path to “reopening” the economy will require strategies for suppressing transmission of SARS-CoV-2 that do not depend exclusively on stringent interventions and such intensive social distancing policies as school and workplace closure and mandatory shelter-in-place (i.e. “lockdowns”). Several different approaches to suppressing transmission have been suggested ([1], [2], [3], [4]), but there has been little systematic comparison of the effectiveness, cost, or robustness of these strategies [5]. We developed models for five approaches to suppressing transmission without the need for completely eliminating personal and business activities. These models illustrate the similarities and differences among these approaches and help to identify their distinctive strengths and weaknesses.

Our conceptual framework distinguishes between *targeted* and *generalized* interventions. Targeted interventions are interventions that are applied to specifically identified individuals in a population, typically based on infection or exposure status. Generalized interventions are behavioral or environmental interventions that are adopted broadly within a population. We consider four targeted interventions that belong to two different strategies that are structurally different in the sense that they are represented by incommensurable flow diagrams.

### Strategy 1: Targeting infected persons

The first strategy targets infected people to limit transmission risk. Each approach in this strategy represents an escalation of intervention.

1. **Active case finding**. Active case finding refers to all efforts that actively seek to identify cases, for instance by testing of health care workers and others who may have high occupational exposures, testing contacts of cases, and adopting minimally exclusive testing criteria. It is assumed that identified cases are isolated and that onward transmission is eliminated or greatly reduced upon isolation. Basically, we are equating active case finding to widespread testing. Active case finding contrasts with passive case finding, which we define as the detection of cases among symptomatic patients who present to medical services for diagnosis of symptoms and receive a test only after meeting some criteria.
2. **Contact tracing**. Contact tracing is the identification, communication with, and monitoring of possible exposures of known cases. Contact tracing increases awareness among the subset of the population most likely to develop symptoms, decreases transmission from traced contacts who are encouraged to isolate, and increases the rate of case finding in the population. Contact tracing may be performed by interviewing cases or family members of cases or with technological aids like cell phone apps [3]. Prior to the 2020 COVID-19 pandemic, contact tracing had never been attempted at the scale that would be required to be effective in suppressing SARS-CoV-2 and several studies have considered how such scale-up might be accomplished [2], [3], [4].
3. **Quarantine**. Quarantine represents an escalation of intervention severity that amplifies the impact of contact tracing. This approach involves isolating traced contacts to the same degree that known cases are isolated. The major effect of this approach is that it reduces the dependence on finding secondary cases (because secondary cases are already identified as contacts) and reduces or eliminates onward transmission from these cases (because the case is already in isolation when symptoms begin). Another effect is that it reduces the average contact rate within the population. Effectively, the portion of the population that is in quarantine is engaged in intensive social distancing, which can be thought of as a “partial lockdown” that is tunable based on the intensity of contact tracing.

### Strategy 2: Targeting uninfected persons

The second strategy comprises one approach targeting healthy people to limit exposure.

1. **Certification**. Certification is an approach that relaxes social distancing in stages. Under this approach, individuals are certified to be infection free before returning to daily routines such as school, work, and shopping. Certification can be *durable* (valid for an extended period of time, for instance based on an antibody test) or *temporary* (valid for a short period of time, for instance because one has recently tested negative by RNA test). Durable certification doesn’t lead to a reduction in transmission, but may be essential for the provision of essential goods and services during periods of high transmission, as conceived by the “shield immunity” concept of Weitz et al. [6].

We note that these strategies have different political, philosophical, ethical and behavioral implications. For instance, Strategy 1 may disincentivize care-seeking because receiving a positive test could preclude one from working whereas Strategy 2 may incentivize care-seeking because a negative diagnostic test or positive antibody test is required to work. Similarly, Strategy 1 prioritizes a right to work whereas Strategy 2 prioritizes a duty to protect. In addition, Strategy 1 and Strategy 2 approaches could be combined. But, because they are structurally different, we do not consider such combinations here.

### Generalized interventions

In addition, these targeted interventions may be used in combination with generalized interventions. Generalized interventions act by reducing transmission or exposure broadly in a population and are not structurally different to the targeted strategies they are combined with in the sense that they may be added to either targeted strategy without modifying the topology of the flow diagram.

1. **Generalized interventions**. Generalized interventions are behavioral or environmental interventions that are adopted broadly within a population, including: wearing face masks [7]; improved hand hygiene [8], [9]; improved cleaning and disinfection of surfaces [9], [10]; greater provision of sick leave and increased enforcement of school and workplace guidelines for staying home when sick [11]; contactless transactions [12]; use of infection barriers in stores, restaurants, and waiting areas; distribution of hand sanitizer in public places; behavioral change (e.g. elbow/fist bump vs. handshake [13]); use of personal rather than public transport; micro-social-distancing (e.g. limiting physical contact, queue spacing); and public policies that limit local aggregations of people such as limits on the number of people allowed in a store and disallowing large events.

### Overview

Below, we present conceptual models devised to be realistic for SARS-CoV-2, but they are not fit to data from any particular population. We studied the dynamics of active case finding, contact tracing, quarantine, and certification individually and in combination with generalized interventions after a “first wave” that infects a small fraction of the population. For comparison, we also consider the two limiting cases of maintaining intensive social distancing and doing nothing. The models are parameterized for a population of 10 million people, slightly larger than London (8.9 million) and New York City (8.3 million) and slightly smaller than the US state of Georgia (10.6 million), but they may be parameterized for a population of any size.

We use the models to answer a number of general strategic questions about these five approaches to suppressing transmission without social distancing.

1. How much might generalized interventions (without targeted interventions) reduce the total outbreak size compared with reference scenarios?
2. When are contact tracing and quarantine most beneficial?
3. What benefit does quarantine add to contact tracing?
4. When can certification be effective?
5. How does the extent of presymptomatic transmission affect the choice of intervention strategy?

## Methods

### Strategy 1: Active case finding, contact tracing, and quarantine

The system of equations for Strategy 1 is

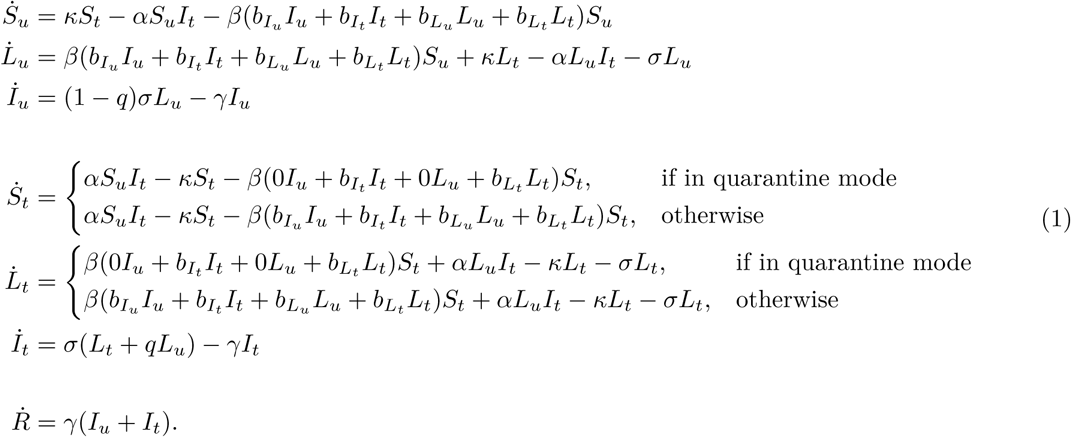

This model (Fig. 1) supposes that there are both traced and untraced persons who are susceptible, incubating, and symptomatic as well as one pool of recovered and deceased, designated *S, L, I*, and *R*. In contrast to the usual convention (where incubating cases are considered to be “exposed”, designated *E*), as presymptomatic transmission is well documented and possibly quite important in the context of COVID-19 interventions ([14], [15], [16]), our model replaces *E* with *L* (for “latent”) which may contribute to the force of infection. Upon displaying symptoms, untraced exposed individuals are assumed to enter isolation with probability *q* ≤ 1, reflecting t he combined effects of both imperfect case ascertainment and contact tracing t hat may include less than 100% of known cases. Incomplete contact tracing could be either intentional or unintentional. It is assumed that all traced exposed individuals remain in the program as new cases upon the development of symptoms. Susceptible and exposed contacts of symptomatic cases are enrolled in the contact tracing program at rate *α*. Traced individuals that do not develop symptoms are released from the program after a period of time 1*/κ*. Transmission is assumed to occur through mass action.

**Figure 1:**
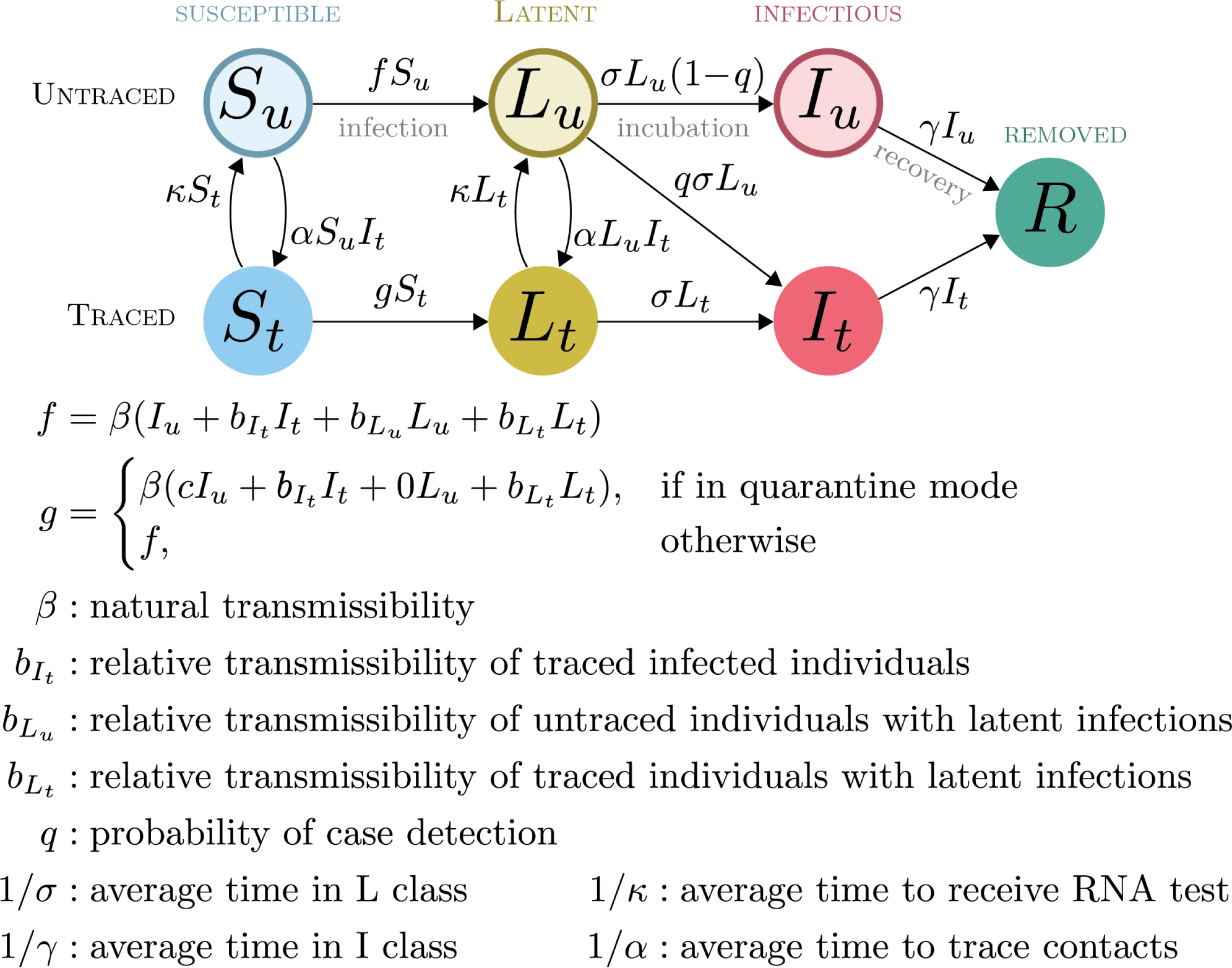
Compartmental model for Strategy 1 interventions.

Using the next-generation matrix ([17], [18], [19]), we obtained the following expression for the basic reproduction number of this model:

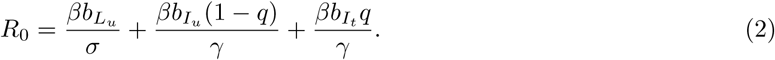

Thus, overall, *R*_0_ is a weighted sum of transmission contributions from the incubating, untraced and traced infectious individuals, respectively. It may sometimes be useful to have the critical case finding value *q*^∗^ at which *R*_0_ = 1. Setting eq 2 to one and solving for *q* obtains

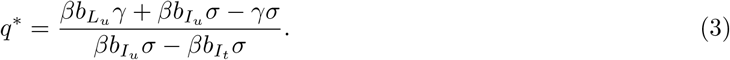

Care should be taken in the interpretation of the force of infection functions *f* and *g*. The force of infection is formulated such that a “natural” transmissibility *β*, assumed to represent the baseline contagiousness of an untraced symptomatic case circulating in the population, is multiplied by a factor 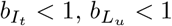 or 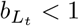 to represent the contagiousness of latent infections and isolated cases. This allows that infection from traced and untraced individuals may occur at different rates and thus we think of the transmissibility “attaching” to the class of the infected individual (traced or untraced). *Completely effective* isolation is represented by setting 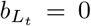 and 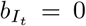. *Active case finding* is represented by setting *α* = 0 and *κ* = 0 and tuning *q* to represent different levels of active case finding. *Quarantine* is represented by setting 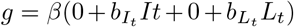 and setting 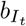 and 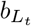 to values that reflect the amount of transmission that may happen within a household where a person is quarantined. Completely effective quarantine is represented by setting 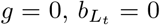 and 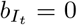. This model reduces to the standard *SEIR* model when 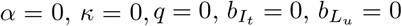, and 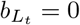. The parameters *q, κ*, and *α* are considered to be control parameters, while the remaining parameters are considered to be natural.

### Strategy 2: Certification

The system of equations for Strategy 2 is

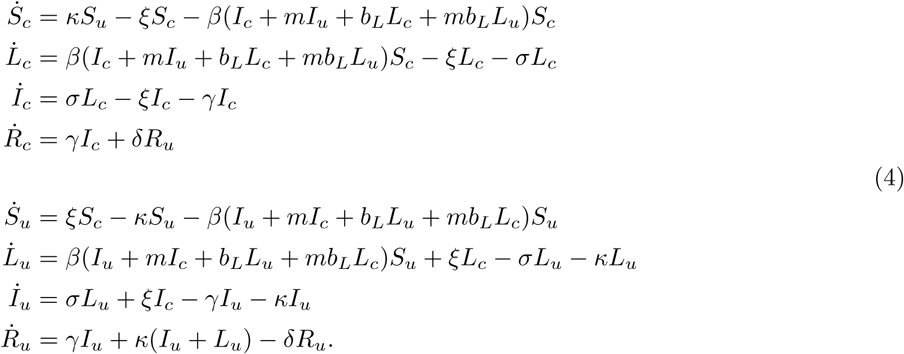

This model (Fig. 2) supposes that there are both certified and uncertified persons who are susceptible, incubating, and symptomatic (designated *S, L*, and *I*), but two pools of removed (*R*). As above, it is assumed that *L* class individuals may contribute to the force of infection. The model allows that infection-free status may be conferred by either serological testing confirming past infection (durable certification) or having recently received a negative RNA test (temporary certification). We suppose that the primary purpose of certification is to change the patterns of contact between certified and uncertified people, i.e. with uncertified individuals practicing intensive social distancing by sheltering in place, not going to school or work, and not participating in large gatherings. In contrast to Strategy 1, here we assumed that *β* attaches to encounters such that encounters within a class (certified-certified or uncertified-uncertified) have one rate of infectious contacts (*β*) while infectious encounters between classes (certified-uncertified) occur at another rate (*βm*), where *m <* 1 is a factor that represents the reduction in mixing. Additionally, factor *b*_*L*_ *<* 1 reduces the infectiousness of incubating infections compared with symptomatic infections. It is assumed that the temporary certification is valid for 1*/ξ* days. The parameters *κ* and *ξ* are considered to be control parameters, while the remaining parameters are considered to be natural.

**Figure 2:**
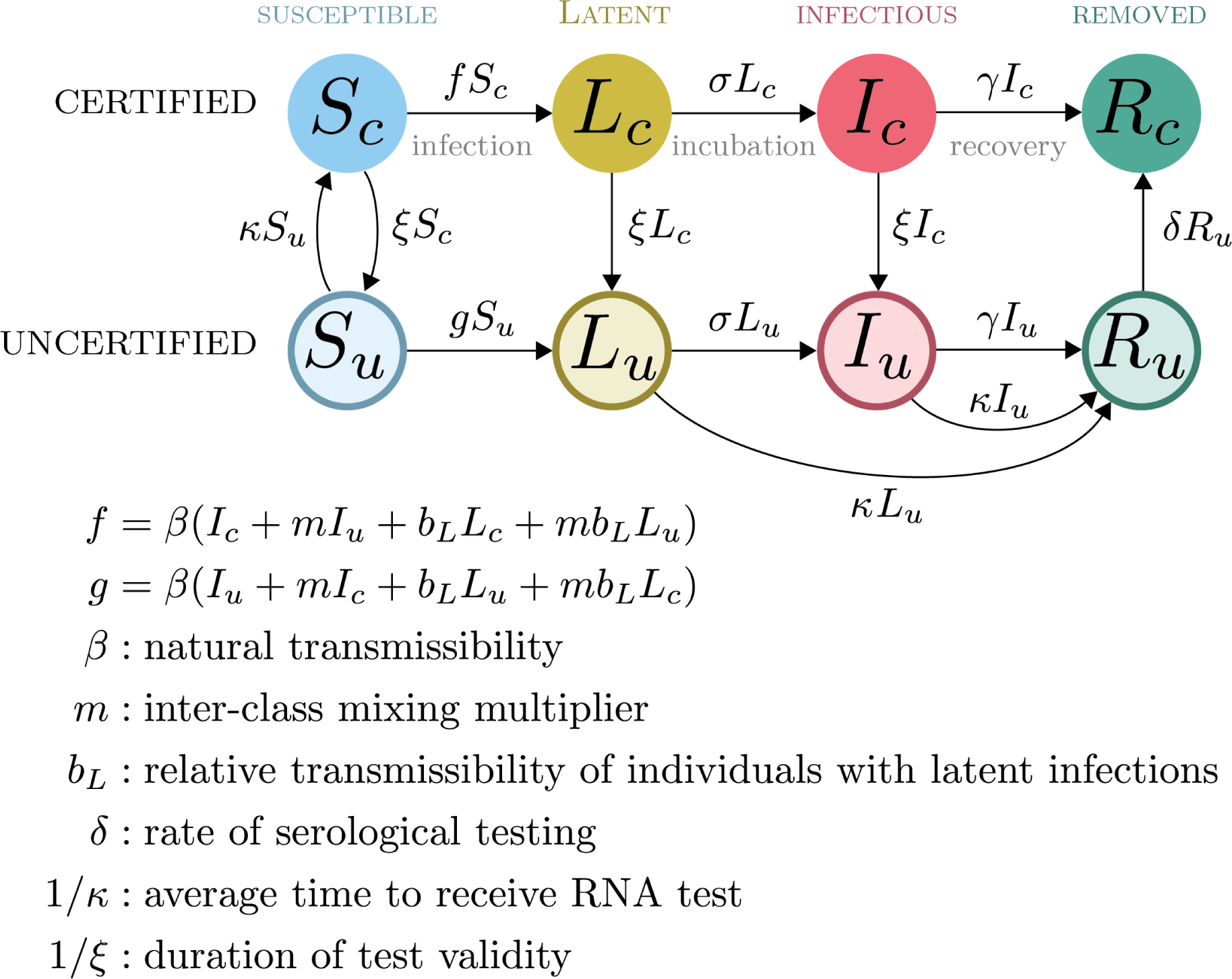
Compartmental model for certifying infection status.

Assuming that there is no certification process prior to the start of the epidemic (i.e. *S*_*c*_ (0) = 0, *S*_*u*_ (0) = 1, and *κ* = 0 initially), then the basic reproduction number for this model is

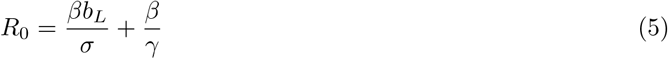

which is the standard form of the basic reproduction number for an SEIR-type compartmental model. If certification is initiated before the start of the epidemic (i.e. if *S*_*c*_ (0) *>* 0, the form of the basic reproduction number is much more complicated. Let *R*_*XY*_ denote the average number of new infections (in compartment *Y*) induced by the introduction of a single individual (in compartment *X*) into a completely susceptible population, over the course of the infectious period of this individual. Then the basic reproduction number takes the form:

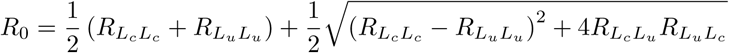

The details of this case and other observations can be found in Appendix 1.

The basic reproduction number is strictly increasing with the inter-class mixing multiplier, *m*. If the classes do not mix at all (*m* = 0), then the basic reproduction number is equal to the greater of the two direct reproduction numbers:

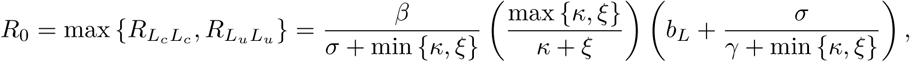

which is in general a lower bound for the value of the basic reproduction number. On the other hand, if the certification process has no impact on the mixing between classes (*m* = 1), then the basic reproduction number will be much larger in general: 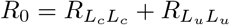. Similarly, this is an effective upperbound for *R*_0_.

Looking at the other control parameters, the basic reproduction number is strictly decreasing in the rate of certification testing, *ξ*. If certification remains valid indefinitely (*ξ* = 0), then the risk of an outbreak is the same as if there were no certification process whatsoever and the basic reproduction number is as above:

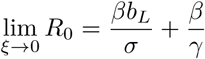

On the other hand, if certification has no effect (*ξ* → *∞*) then there is a significantly lower risk of an outbreak:

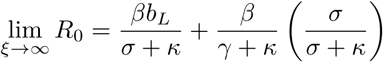

Hence, without certification, the ability to prevent an outbreak is determined solely by the rate of RNA testing (*κ*).

We obtain similar results for the limiting cases for the rate of RNA testing (*κ*):

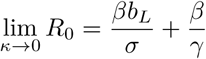

and

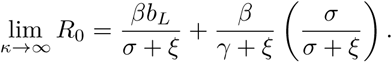

### Generalized interventions

Because transmission is the result of contagious contact, targeted and generalized interactions have multiplicative effects. In our model, generalized interventions are represented by multiplying *β* by a factor less than one (typically 0.7, corresponding to a 30% reduction in transmission).

### Implementation

Solutions to the equations were obtained using the R package pomp [20]. The fundamental transmission parameters are assumed to be *β* = 0.5 and *γ* = 1*/*6 so that the basic reproduction number *R*_0_ = *β/γ* = 3.0. We assume an incubation period of four days (*σ* = 1*/*4). We assume that incubating infections are 30% as contagious as symptomatic infections and that isolation reduces transmission by 90% so 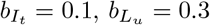, and 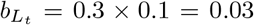. For comparability between Strategy 1 and Strategy 2, we assume that the reduction in mixing due to certification is similar to the reduction in transmission due to isolation and set 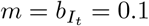. Social distancing is assumed to reduce transmission by 60% while generalized interventions are assumed to reduce transmission by 30%.

Typically, we study the sensitivity of the final epidemic size to the choice of control parameters *q, α, κ, ξ*, and *δ*, but consider the values *q* = 0.5, *α* = 10 × *β* = 5, *κ* = 1*/*3, *ξ* = 1*/*7, and *δ* = 1*/*10 as a reference point, implying case finding of 50%, that five contacts are traced for every secondary infection, that the delay to obtain a diagnostic test is three days, that diagnostic certification is valid for seven days, and that the time to obtain an antibody test is 10 days. For comparison, we note that the CDC considers 50% to be the upper bound on the percentage of cases that are asymptomatic ([10], [21]). The sensitivity of our conclusions to these choices is studied in greater detail in Appendix 2. We assume transmission is initiated with 1,000 infected individuals evenly distributed between incubating and symptomatic compartments of the non-target class (i.e. untraced or uncertified).

## Results

### How much might generalized interventions (without targeted interventions) reduce the total outbreak size compared with reference scenarios?

The reference condition of continued social distancing is represented in the certification model by setting *ξ* and *κ* to 0 and setting *β*_0_ to 40% of its original value. (For comparison, equivalent baseline conditions for the contact tracing model are provided in Appendix 2.) At the assumed level of social distancing, a large outbreak still occurs, ultimately infecting approximately half of the population (Fig. 3, top). Social distancing combined with generalized interventions does not result in complete suppression, but reduces transmission to very close to the critical level.

**Figure 3:**
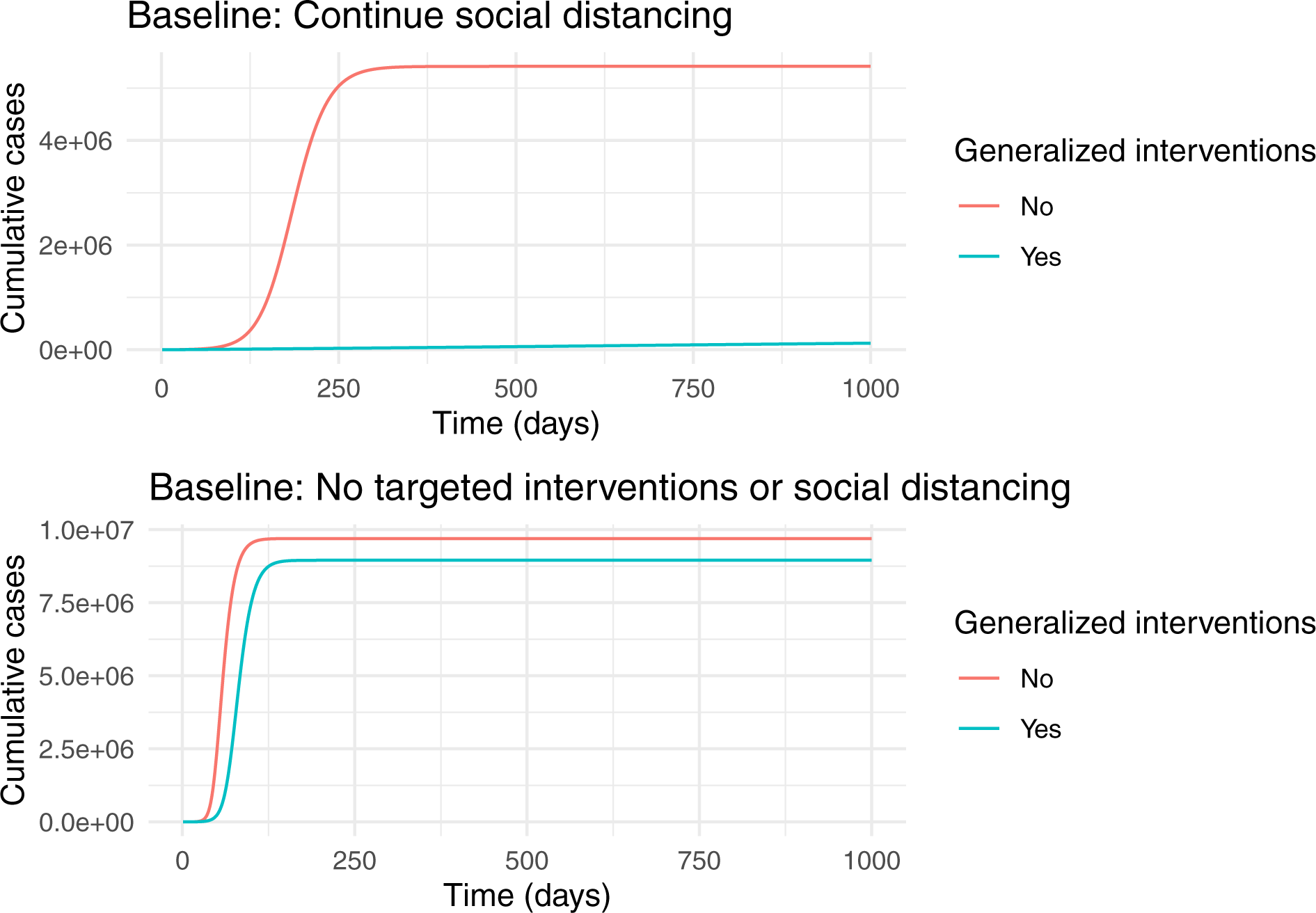
Two baseline scenarios. The top plot assumes that transmissibility, *β*, is at 40% of its natural value. The bottom plot assumes that transmissibility, *β*, is at its natural value (*β* = 0.5). Both plots assume that generalized interventions reduce transmissibility by a further 30%. Other parameters are *b*_*L*_ = 0.3, *m* = 0.1, *γ* = 1*/*6, *σ* = 1*/*4, *κ* = 0, *δ* = 1*/*10, and *ξ* = 1*/*7.

A scenario with no social distancing and no targeted interventions is represented by setting *ξ* and *κ* to 0 and *m* = 1. Unsurprisingly, the large majority of the population is infected under this condition and generalized interventions only reduce the total outbreak size by a relatively small amount (Fig. 3, bottom). These results suggest that generalized interventions of the magnitude envisioned here are not sufficient to suppress transmission. If continued social distancing is not possible, then targeted interventions will be essential if infection of the majority of the population is to be prevented.

### When are contact tracing and quarantine most beneficial?

Active case finding, contact tracing and quarantine represent an escalation of Strategy 1 approaches to suppressing transmission. As a baseline, it is therefore useful to understand the conditions, if any, under which active case finding alone can limit transmission. To investigate active case finding as a control parameter, we set *α* = 0 and *κ* = 0 and plot the final epidemic size as a function of *q*. Complete suppression without generalized interventions requires case finding to identify approximately 95% of cases (Fig. 4, green line).

**Figure 4:**
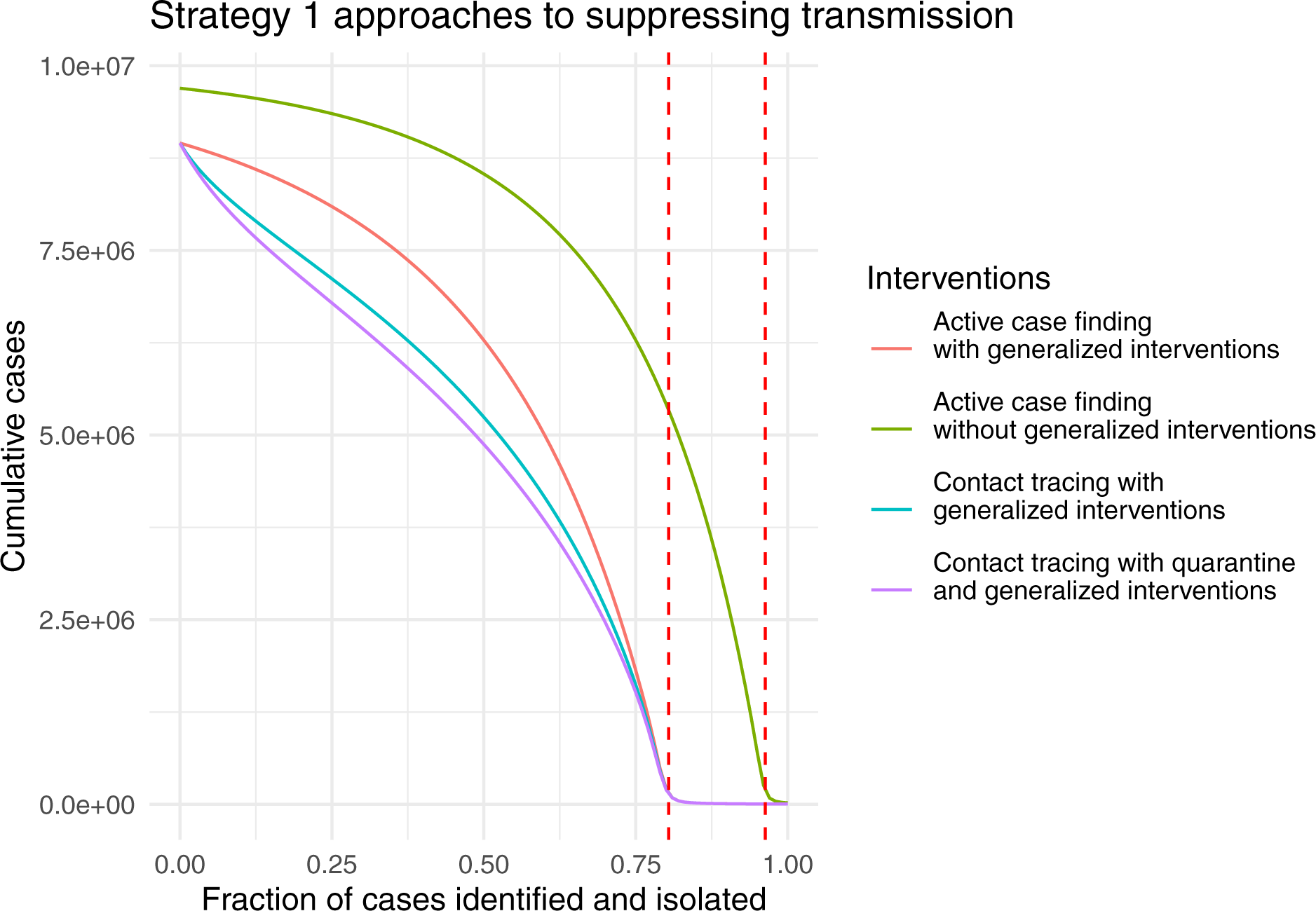
Strategy 1 approaches to suppressing COVID-19 transmission as a function of case ascertainment (*q*). Dashed red lines show the critical value *q*^∗^ at which *R*_0_ = 1 with and without generalized interventions. Other parameters are 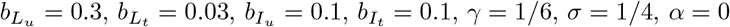, and *κ* = 0.

This seems untenable for a disease that is symptomatic in only approximately 80% of cases. The addition of generalized interventions reduces the critical value for case finding to around 80% (Fig. 4, red line), which still seems like a great challenge. At a more realistic level of 50% case finding, greater than half of the population would be infected with generalized interventions and around 80% without generalized interventions. For comparison, many scientists and health experts think case ascertainment of COVID-19 in a number of settings was originally between 1% and 10% ([22], [23]), so 50% represents finding about five times as many cases as occurred during the first wave. It seems implausible that 50% case finding could occur without widespread testing. We also show the relative impact of contact tracing and quarantine (Fig. 4, blue and purple lines). For parameters studied here, the relative additional benefit provided by quarantine is quite small compared with contact tracing. Further, contact tracing and quarantine do not change the value of case finding at which suppression is achieved, but do reduce the total number of cases for a given level of case finding below the critical value of *q*^∗^. The relative additional benefit obtained by contact tracing and quarantine is at its maximum at relatively small values of case finding around 25%.

### What benefit does quarantine add to contact tracing?

These results are possibly surprising. Particularly, why isn’t quarantine more effective compared with contact tracing, given that it has been such a longstanding public health strategy? Our model assumes that quarantined individuals are excluded from encounters in the general population. But, in recognition that traced contacts will often be family members and expecting that family members may be quarantined together, the model allows for transmission at 10% of the baseline value. We wondered if this small amountof transmission from quarantined individuals to family members accounts for the difference. To investigate this idea, we repeated the analysis setting 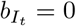 and 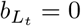, turning off transmission to or from traced contacts entirely. The overall shape of the effect of case identification on total outbreak size is similar, but shifted (Fig. 5).

**Figure 5:**
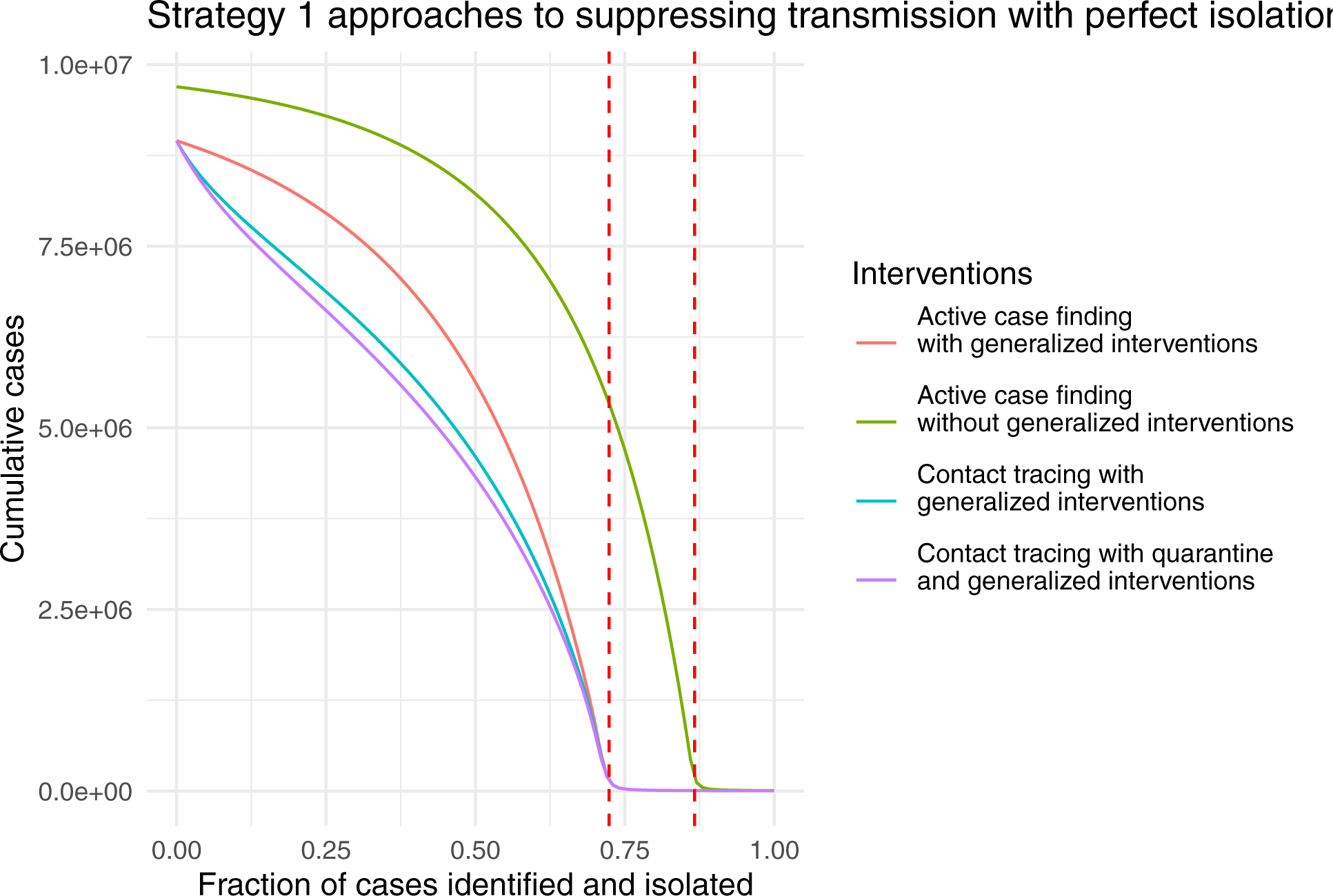
Strategy 1 approaches to suppressing COVID-19 transmission as a function of case ascertainment (*q*) with perfect isolation. Dashed red lines show the critical value *q*^∗^ at which *R*_0_ = 1 with and without generalized interventions. Other parameters are 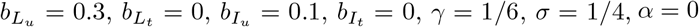, *γ* = 1*/*6, *σ* = 1*/*4, *α* = 0, and *κ* = 0.

By comparing the curves in Figures 4 and 5, we see that eliminating this last 10% of transmission increases the total number of cases averted approximately tenfold from 250,000 to almost 2,500,000, over a large range of *q* for all three Strategy 1 approaches (Fig. 6).

**Figure 6:**
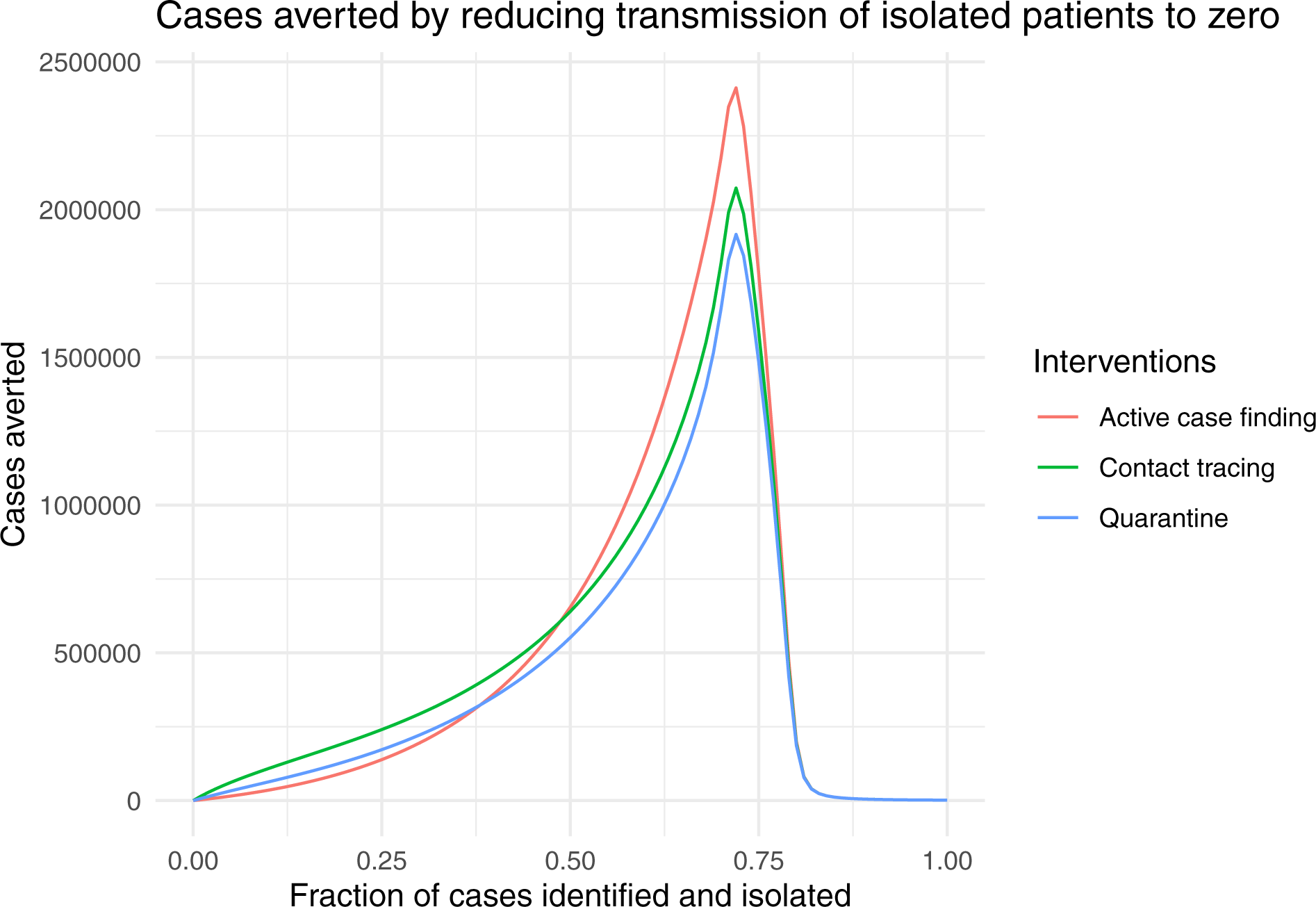
Cases averted by reducing transmission from isolated patients from 10% to zero (*b*_*L*_ = 0, *b*_*I*_ = 0) as a function of case ascertainment (*q*). Other parameters are 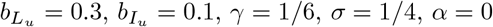, and *κ* = 0.

### When can certification be effective?

Here we look at certification with and without generalized interventions. To better understand the range of conditions under which certification can be effective, we examine the final outbreak size over a grid comprising all combinations of viral test validity from 7 to 21 days and for test waiting times from one to five days (Fig. 7). Interestingly, in both cases there is a very sharp boundary between those testing regimes in which suppression of transmission is achieved (dark blue) and testing regimes where a very large outbreak ensues. These results suggest that it is virtually impossible to suppress transmission without generalized interventions. The “safe” region (dark blue) is larger when there are generalized interventions (Fig. 8). Specifically, it appears that a test validity of 7-10 days together with a waiting time of no more than 3 days would achieve suppression. However, the sharpness of the boundary between suppression and a failure to suppress suggests that this approach is fragile, such that small inaccuracies in parameter values or model specification maycause the approach to fail.

**Figure 7:**
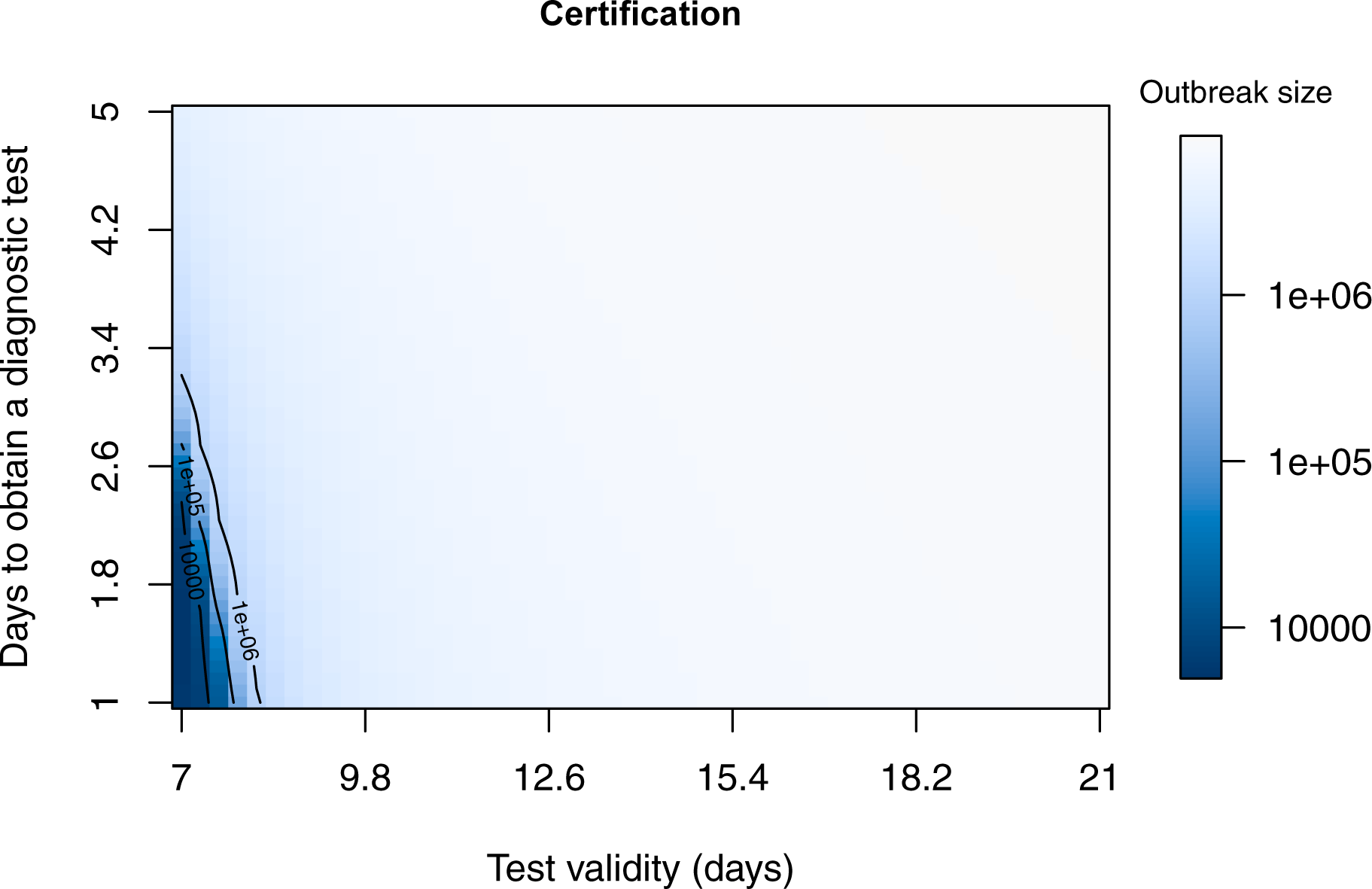
Final outbreak size as a function of viral test validity (1*/ξ*) and test lag (1*/κ*) without generalized interventions. At the assumed value of presymptomatic transmission (*b*_*L*_ = 0.3), there is only a very small region (dark blue) within which certification can prevent a major epidemic. Other parameters are *β* = 0.5, *m* = 0.1, *γ* = 1*/*6, *σ* = 1*/*4, and *δ* = 0.1.

**Figure 8:**
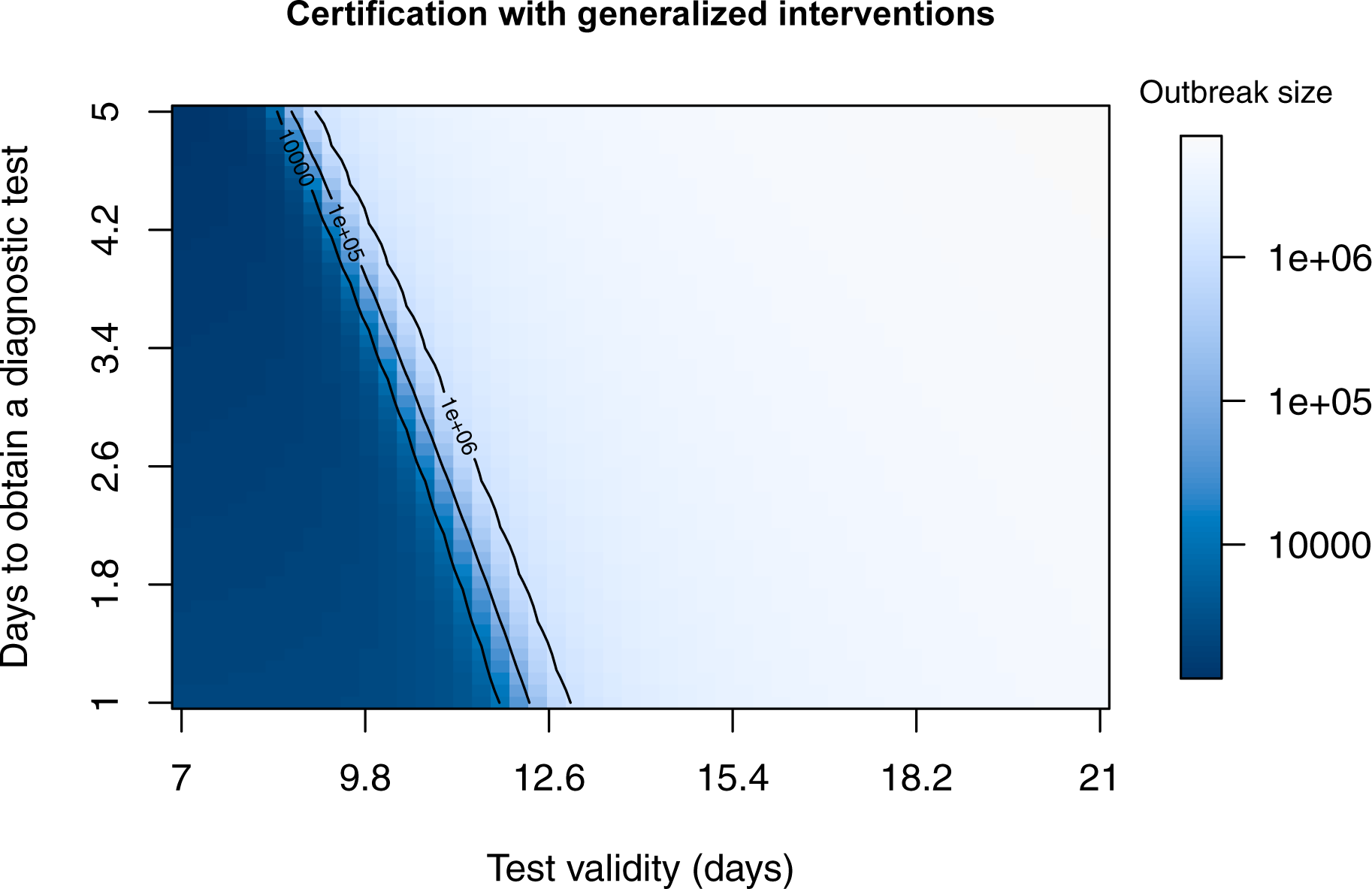
Final outbreak size as a function of test validity (1*/ξ*) and test lag (1*/κ*) with generalized interventions. At the assumed value of presymptomatic transmission (*b*_*L*_ = 0.3), there is modest region (dark blue) within which certification can prevent a major epidemic. Other parameters are *β* = 0.5, *m* = 0.1, *γ* = 1*/*6, *σ* = 1*/*4, and *δ* = 0.1.

### How does the extent of presymptomatic transmission affect the choice of intervention strategy?

The preceding analyses assume that latent cases are 30% as infectious as symptomatic cases, but it is well known that “silent transmission” is a key component of COVID-19 epidemiology [14] [15]. Here we investigate how different levels of presymptomatic transmission influence the effectiveness of the containment strategies introduced here. First, we plot the total outbreak size against the assumed level of infectivity (i.e. the parameter 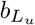; for each level of 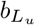, the transmissibility of traced individuals is set to 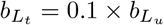 (Fig. 9).

**Figure 9:**
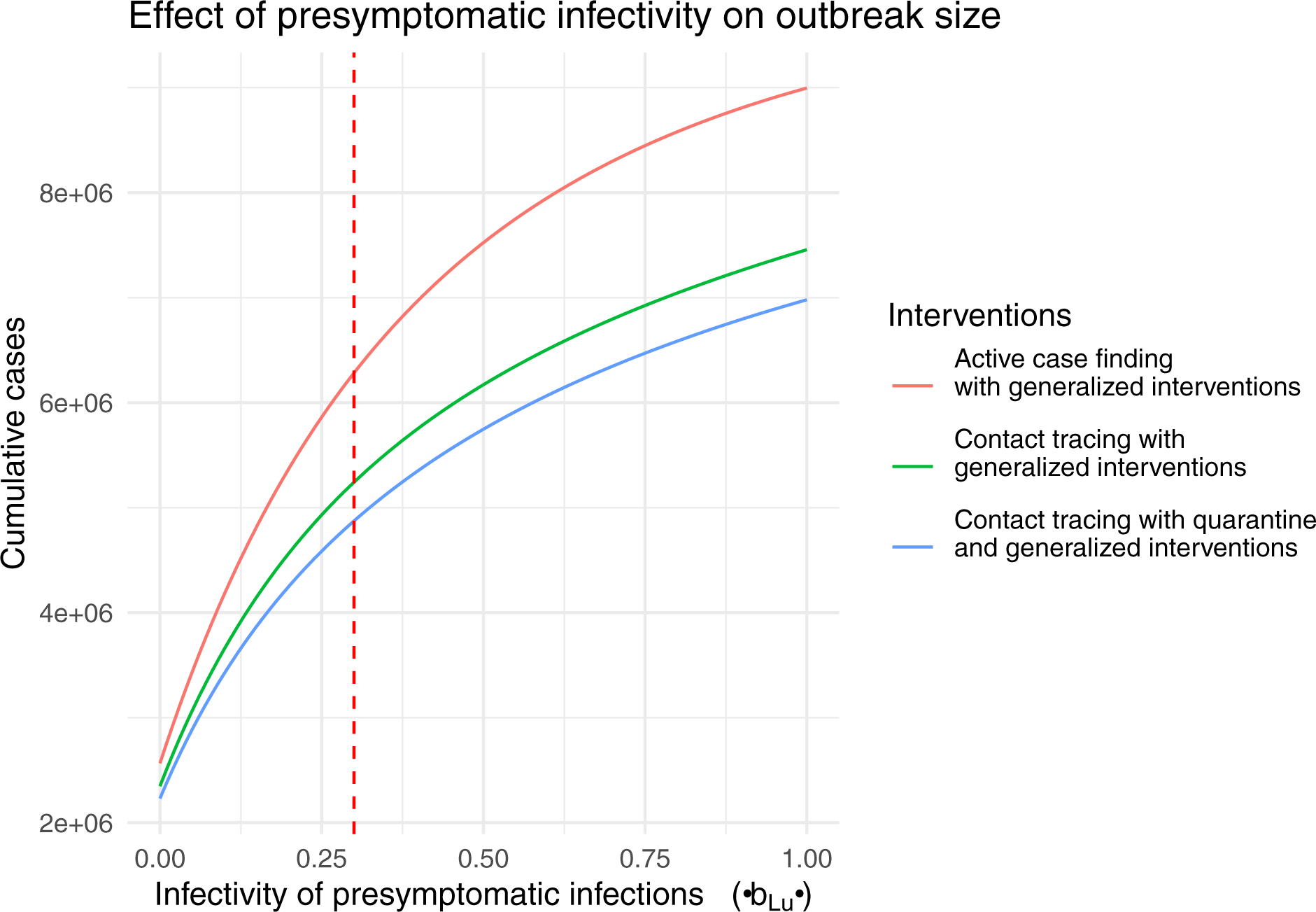
Effect of presymptomatic infectivity on outbreak size for 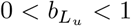. The vertical dashed line shows the default value of 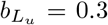 for comparison with other figures. Other parameters are 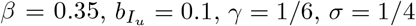, and *q* = 0.5.

Next we look at the certification model at four different levels of *b*_*L*_. Epidemic outcomes are summarized by plotting the contour for combinations of test validity (1*/ξ*) and test lag (1*/κ*) where the final outbreak size is 10,000 (Fig. 10). Because the transition is so sharp (compare Fig. 8), this is effectively the “containment boundary” separating minor transmission and a major epidemic. Unsurprisingly, for presymptomatic transmission less then the default value of 0.3, a longer test validity and test lag may be tolerated without risking a major outbreak. However, even with no presymptomatic transmission (0% contour), the safe region remains relatively small with a maximum test validity of around two weeks. As presymptomatic transmission approaches the level of symptomatic transmission, the safe region dimininishes substantially.

**Figure 10:**
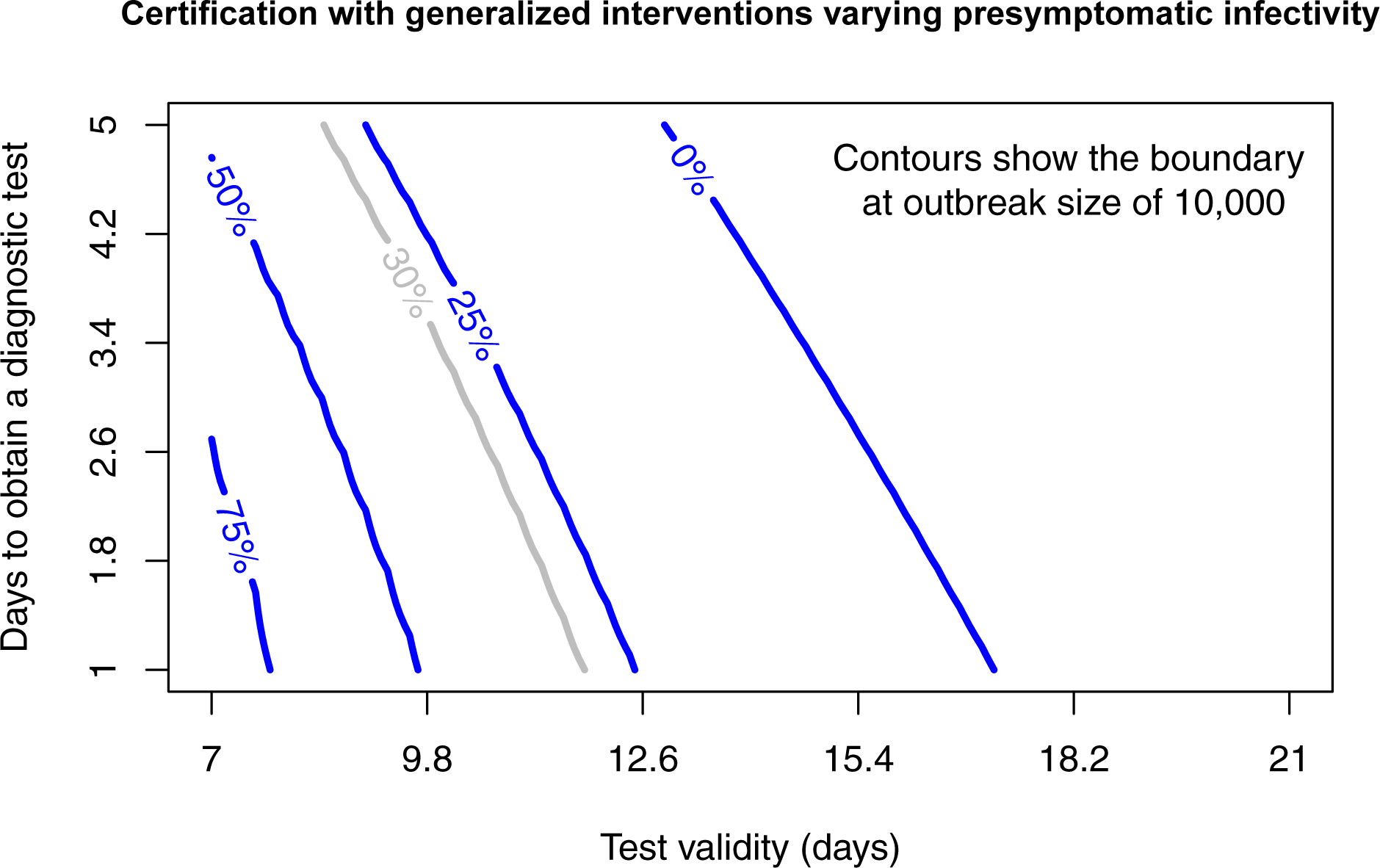
Certification with generalized interventions for presymptomatic infectivity (*b*_*L*_) assumed to be 0%, 25%, 50%, and 75% of the baseline value of *β* = 0.35 for transmission with generalized interventions. Combinations of test validity and days to obtain a test that are below the contour have total outbreak sizes less than 10,000 and may be considered to be “contained”. The default value of *b*_*L*_ = 0.3 is plotted in grey for comparison with Figure 8. Other parameters are *m* = 0.1, *γ* = 1*/*6, *σ* = 1*/*4, and *δ* = 0.1.

## Discussion

### Are our parameters realistic for COVID-19?

In most of the scenarios studied, we assumed that active case finding would yield case ascertainment rates of 50%. For context, this can be compared with either estimated case ascertainment rates or estimated symptomatic rates (which sets an upper bound on case acertainment through clinical diagnosis). In analysis of data from passengers on the Diamond Princess cruise ship, Mizumoto et al. [24] estimated an overall asymptomatic proportion of 17.9% (equating to a symptomatic proportion of 82.1%). Among residents in a nursing home, 10 out of 23 (43.5%) were symptomatic at the time of testing [25]. A review of multiple populations finds that the fraction of asymptomatic persons infected with SARS-CoV-2 may be 45-50% [21]. For comparison, estimates of ascertainment in the US in for Spring 2020 are in the range of 1-10% ([22], [23]).

### Strategic approaches to suppressing transmission

Here we have analyzed the two structurally different approaches to suppressing transmission without intensive social distancing (i.e. “lockdowns”). These two strategies are not directly comparable in the sense that they are represented by incommensurable flow diagrams (Figs. 1 and 2) and neither is a special or limiting case of the other. Of course, these strategies could be used together for greater effectiveness. But, a flow model to evaluate the optimal use of strategies in combination would be considerably more complicated, requiring approximately 16 states to represent the possible combinations of certified and uncertified persons that may be either traced or untraced and in one of the four primary infection states (*S, L, I*, and *R*). Even though the actual number of flows among these 16 states will be considerably fewer than the 16 × 15 = 240 possibilities, it would be a considerable challenge to sensibly parameterize such a model. Developing such a model could nonetheless be a useful future step toward developing a complete understanding of transmission reduction via non-pharamceutical interventions for acute infectious diseases.

## Conclusions

These results suggest that any of the preceding strategies may suppress transmission, but that suppression depends on achieving a certain level of effectiveness (reduction in transmission among isolated persons, intensity of contact tracing, frequency of certification, etc.) that varies according to the strategy. Particularly, Strategy 1 approaches (active case finding, contact tracing, and quarantine) are expected to work only when case ascertainment is high. In contrast, Strategy 2 approaches (certification) are only expected to work in a narrow range of conditions (i.e. high frequency testing). These findings suggest that, regardless of whether Strategy 1 approaches or Strategy 2 approaches are adopted, a large testing capacity is required and success will depend on the effectiveness of generalized interventions. Additionally, generalized interventions function as a “force multiplier”. In most scenarios we consider to be realistic, generalized interventions will be essential to achieve suppression.

## Data Availability

This study reports no new data

## Appendix 1 *R*_0_ when certification is initiated before the start of the epidemic

Here we provide a description of the basic reproduction number of Strategy 2 in the general case that certification is initiated before the start of the epidemic.

### Disease-free equilibria

In the absence of infections (that is, *L*_*c*_ = *I*_*c*_ = *L*_*u*_ = *I*_*u*_ = 0), at equilibrium the remaining state variables satisfy the equations *ξS*_*c*_ = *κS*_*u*_ and *R*_*u*_ = 0, with *R*_*c*_ arbitrary. The possible disease-free equilibria lie on a line described by 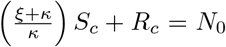, where *N*_0_ is the initial total population size. Assuming that the population of recovered or removed certified individuals is zero (*R*_*c*_ = 0) and that *N*_0_ = 1, we obtain 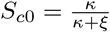 and 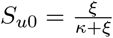. The disease-free equilibrium is then (*S*_*c*0_, 0, 0, 0, *S*_*u*0_, 0, 0, 0).

### Basic reproduction number

We use the method of van den Driessche and Watmough to determine the basic reproduction number as the spectral radius of the next-generation matrix [17]. The model system of equations meets the assumptions necessary to apply this method.

We re-order the state variables so that the infected compartments appear first and then they are sorted by certification status then infection status: *x* = (*L*_*c*_, *I*_*c*_, *L*_*u*,_ *I*_*u*_, *S*_*c*_, *S*_*u*_, *R*_*c*_, *R*_*u*_). The disease-free equilibrium is re-defined to be *x*_0_ = (0, 0, 0, 0, *S*_*c*0_, *S*_*u*0_, 0, 0). The system of ordinary differential equations is decomposed as 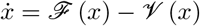 where ℱ (*x*) contains the rate of new infections in each compartment and 𝒱(*x*) the net rates out of each compartment:

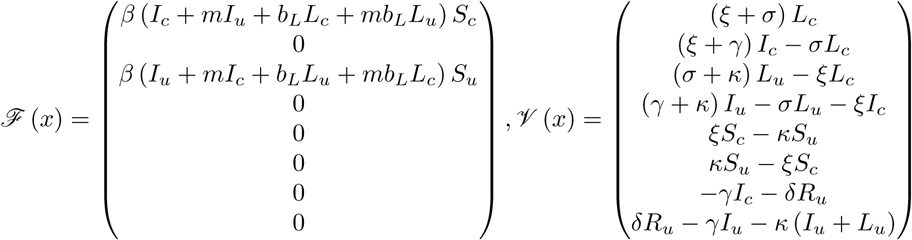

Denoting *F* = [*D* ℱ (*x*_0_)]_1_ ≤_*i,j*_ ≤ _4_ and *V* = [*D*𝒱 (*x*_0_)]_1_ ≤_*i,j*_ ≤_4_, the next generation matrix is defined a s *K* = *F V* ^−1^. In order to better describe the epidemiological meaning of the matrices *F* and *V* ^−1^ in this context, we relabel their entries. The entries of the matrix *F* correspond to the relative contributions of each infectious compartment to the rate of new infections. For example, the components of the first r ow c orrespond to the rate of new infections in the certified classes (i.e. *L*_*c*_) a nd t he c omponents o f t he t hird r ow similarly correspond to new infections in the uncertified classes (i.e. *L* _*u*_). The columns correspond to the compartments contributing to the production of new infections: column 1 is *L*_*c*_, column 2 is *I*_*c*_, column 3 is *L*_*u*_, and column 4 is *L*_*c*_. To make these associations explicit, let *f*_*XY*_ denote the rate of new infections into compartment *Y* induced by individuals in compartment *X*. Then *F* can be written equivalently as

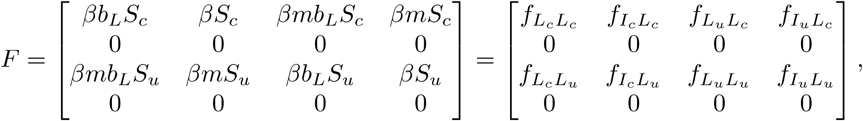

The entries of the matrix *V* ^−1^ correspond to the average amount of time spent in each infectious compartment conditioned on the probability of transitioning from the appropriate previous compartment. Interpretation of the components of *V* ^−1^ is aided by defining transition probabilities between compartments. We use the notation *p*_*XY*_ to refer to the probability of transitioning from compartment *X* to compartment *Y* (before transitioning to other connected compartments). The probabilities appearing in the entries of *V* ^−1^ are

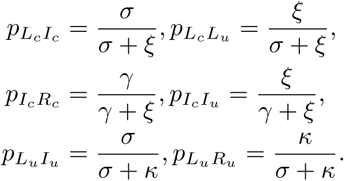

Let *λ*_*X*_ denote the average amount of time spent in compartment *X*. Then *V* ^−1^has equivalent forms:

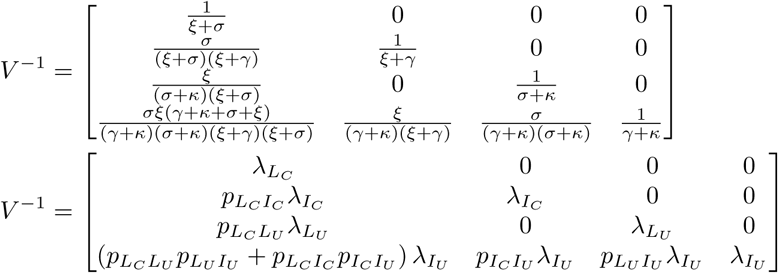

Note that the sum in row four, column one takes into account that there are two paths an individual can take from *L*_*c*_ to *I*_*u*_, depending on whether their certification ends before they become infectious: *L*_*c*_ → *I*_*c*_ → *I*_*u*_ or *L*_*c*_ → *L*_*u*_ → *I*_*u*_.

The next generation matrix, *K* = *FV* ^−1^, is thus given by

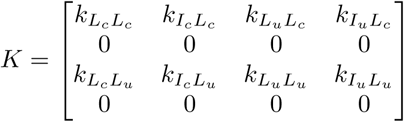

which has non-zero components of:

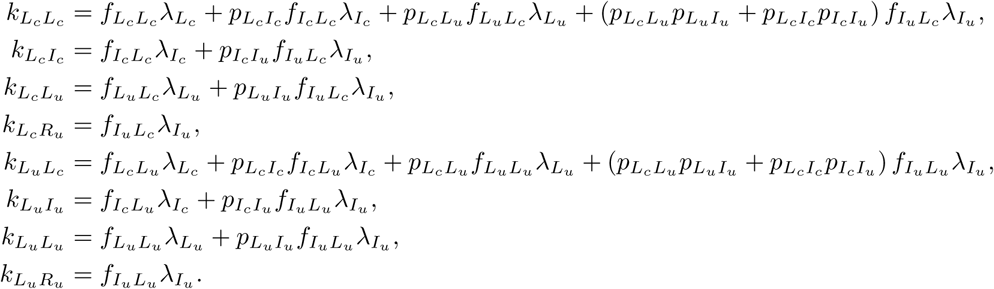

In fact, these quantities can themselves be interpreted as type reproduction numbers. Denote *k*_*XY*_ = *R*_*XY*_, which represents the average number of new infections in compartment *Y* induced by the introduction of a single individual in compartment *X* into a completely susceptible population over the course of the infectious period of this individual. Generally, these are formed from a sum of quantities of the form “Pr (transitioning into compartment *X*) × (rate of new infections in compartment *X*) × (average time spent in compartment *X*).”

Finally, the basic reproduction number *R*_0_ is the largest eigenvalue of the next-generation matrix:

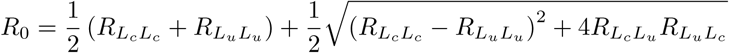

The model admits four type reproduction numbers which are constituent elements of the formula for the basic reproduction number. We use the symbol *R*_*XY*_ to represent the average number of new infections in compartment *Y* induced by the introduction (into a completely susceptible population) of a single infectious individual in compartment *X*, over the course of the infectious period of this individual. Then the general basic reproduction number is given by

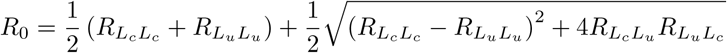

### Observations about the basic reproduction number

We can determine bounds of *R*_0_ :

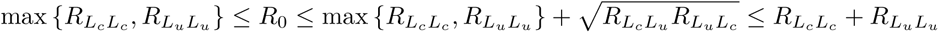

In fact, *R*_0_ is strictly increasing in *m*, the inter-class mixing multiplier. It is equal to its lower bound when there is no mixing between the classes (*m* = 0). On the other hand, when the certification process has no effect on the mixing of the two classes (*m* = 1), *R*_0_ is equal to its largest upper bound. These two cases are elaborated below.

If there is perfect separation between the certified a nd u ncertified classes (*m* = 0), th e ba sic reproduction number meets its lower bound: 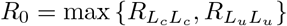 where in this case:

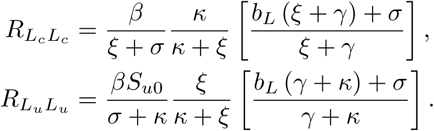

Thus if *m* = 0 the basic reproduction number takes on the value of the larger direct type reproduction number, which is determined by which of *κ* or *ξ* is larger:

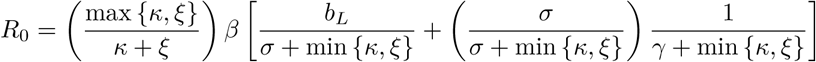

On the other hand, if there is no separation between the certified and uncertified classes (*m* = 1), then 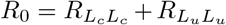. As one might expect, if certification does not lead to a meaningful separation of classes, then the effective contact rate, and hence the basic reproduction number, of the population will be larger. This relation occurs because when 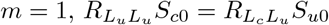 and 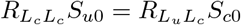.

The parameter *δ*, the rate of serological testing, has no effect on the basic reproduction number.

## Appendix 2 Sensitivity analysis

Here we provide additional details and analysis of models reported on in this paper.

### Baseline scenarios

For comparison, we first plot some baseline scenarios.

#### Baseline 1: Continue social distancing

Continued social distancing is represented by the certification model setting *ξ* and *κ* to 0 and setting *β* to 30% of its original value. Clearly, social distancing will be ineffective without also adopting generalized interventions.

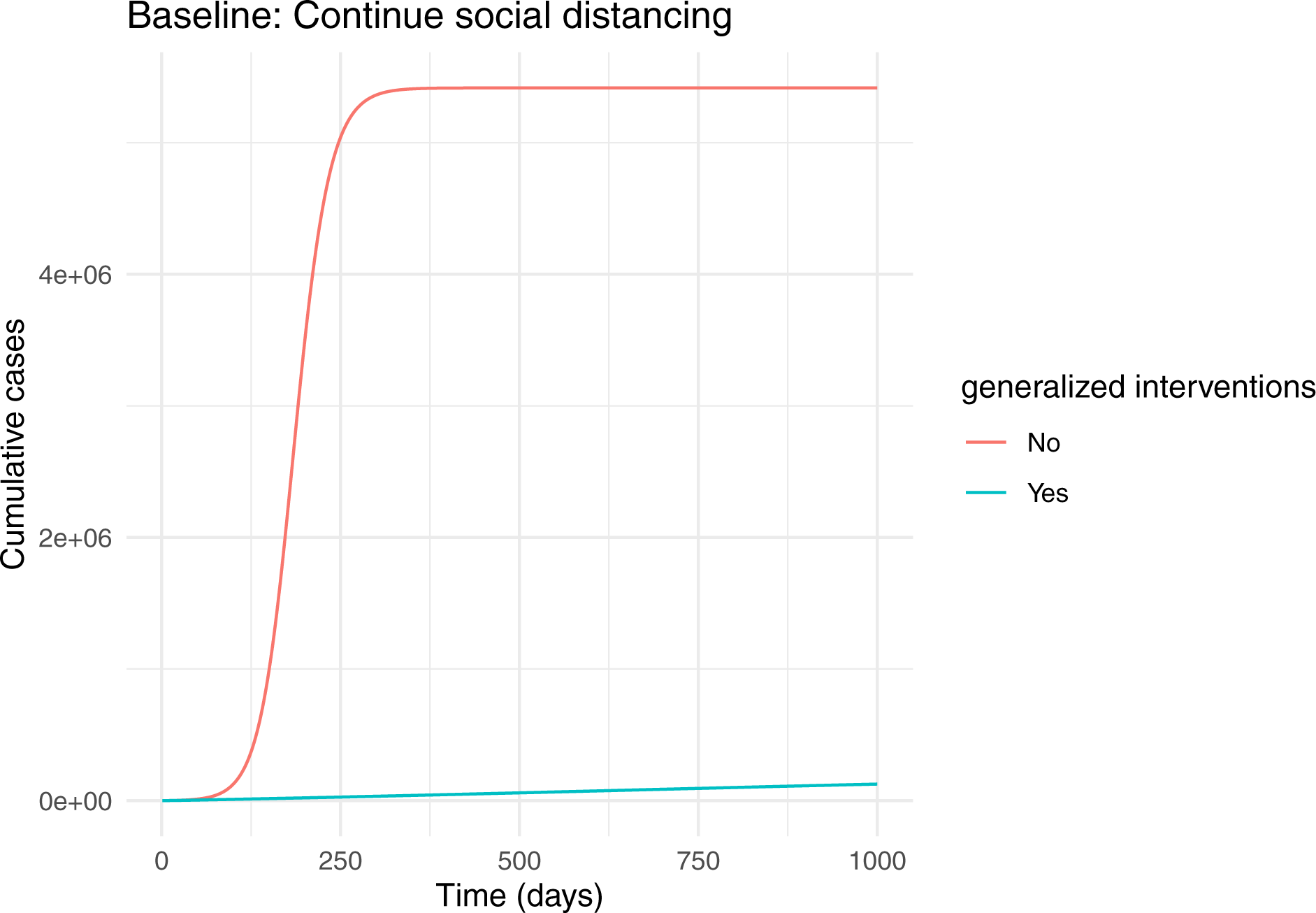

#### Baseline 2: Do nothing

A return to normal is represented by the certification model setting *ξ* and *κ* to 0 and setting *m* = 1.

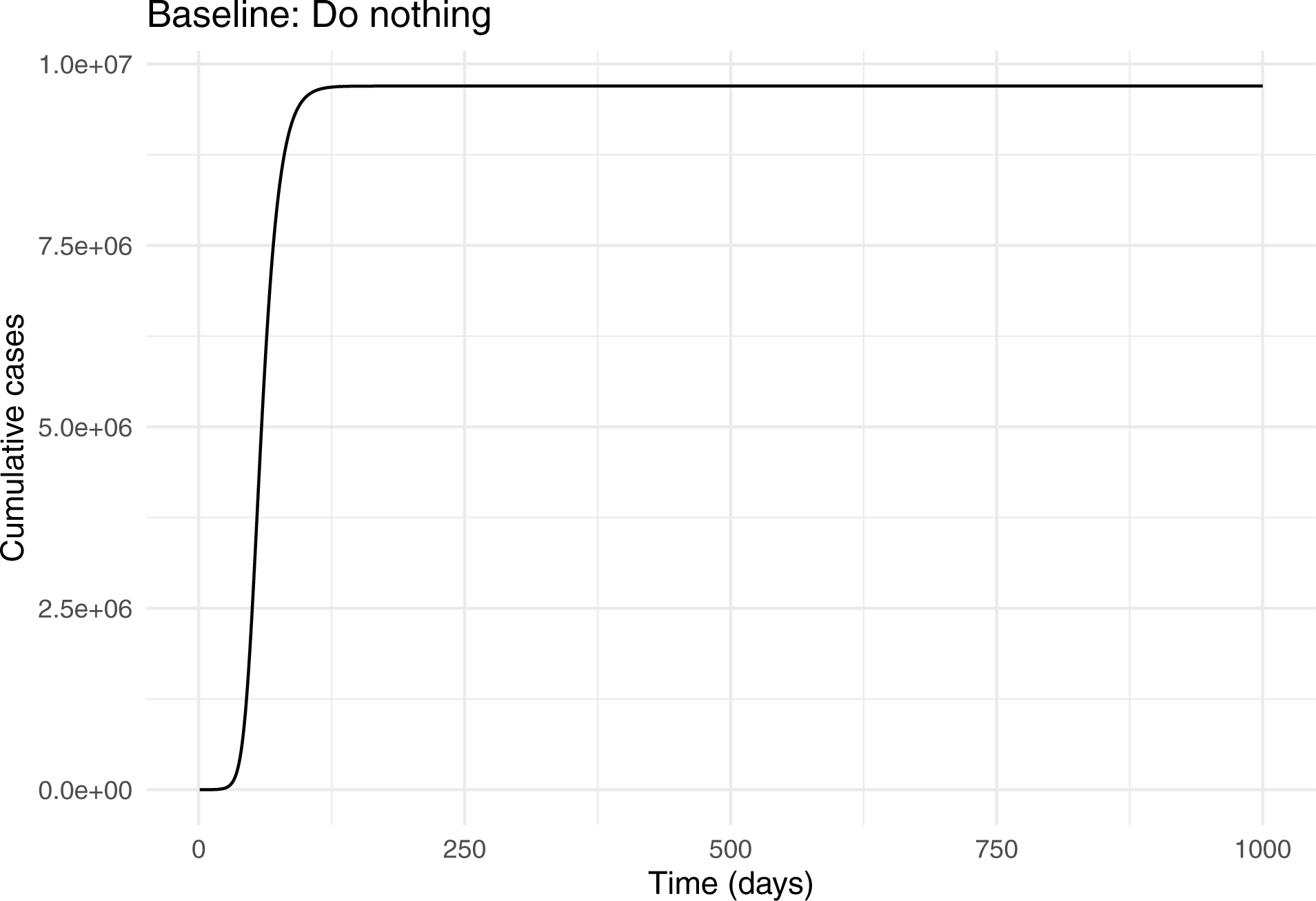

### Scenarios for suppressing transmission without social distancing

#### Active case finding

Active case finding (without contact tracing) is represented by the contact tracing model with *α* = 0, *κ* = 0, and *q* = 0.8. Only *q* is effectively a control parameter for active case finding.

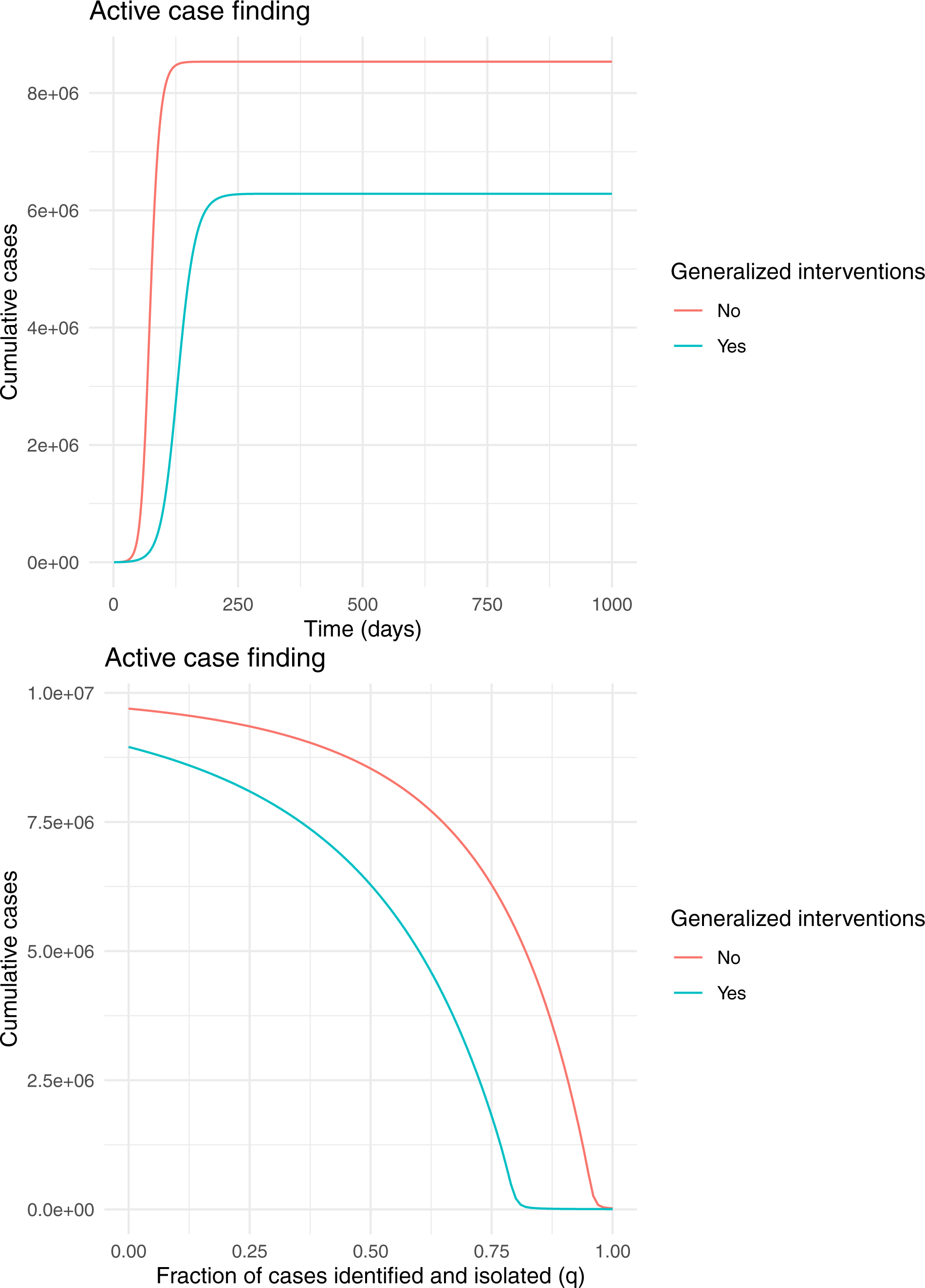

### Contact tracing

For the contact tracing model, we assume that 10 contacts occurred for every one that resulted in transmission (*α* = 10 × *β*_0_) and that contacts are followed for two weeks (*κ* = 1*/*14). This is equivalent to adding contact tracing to the active case finding scenario. We also plot heat maps to examine the sensitivity of outbreak size to the tracing rate and testing intensity in scenarios with and without generalized interventions. As expected, generalized interventions substantially reduce outbreak size. However, outbreak size is relatively insensitive to other control parameters over a reasonably large range.

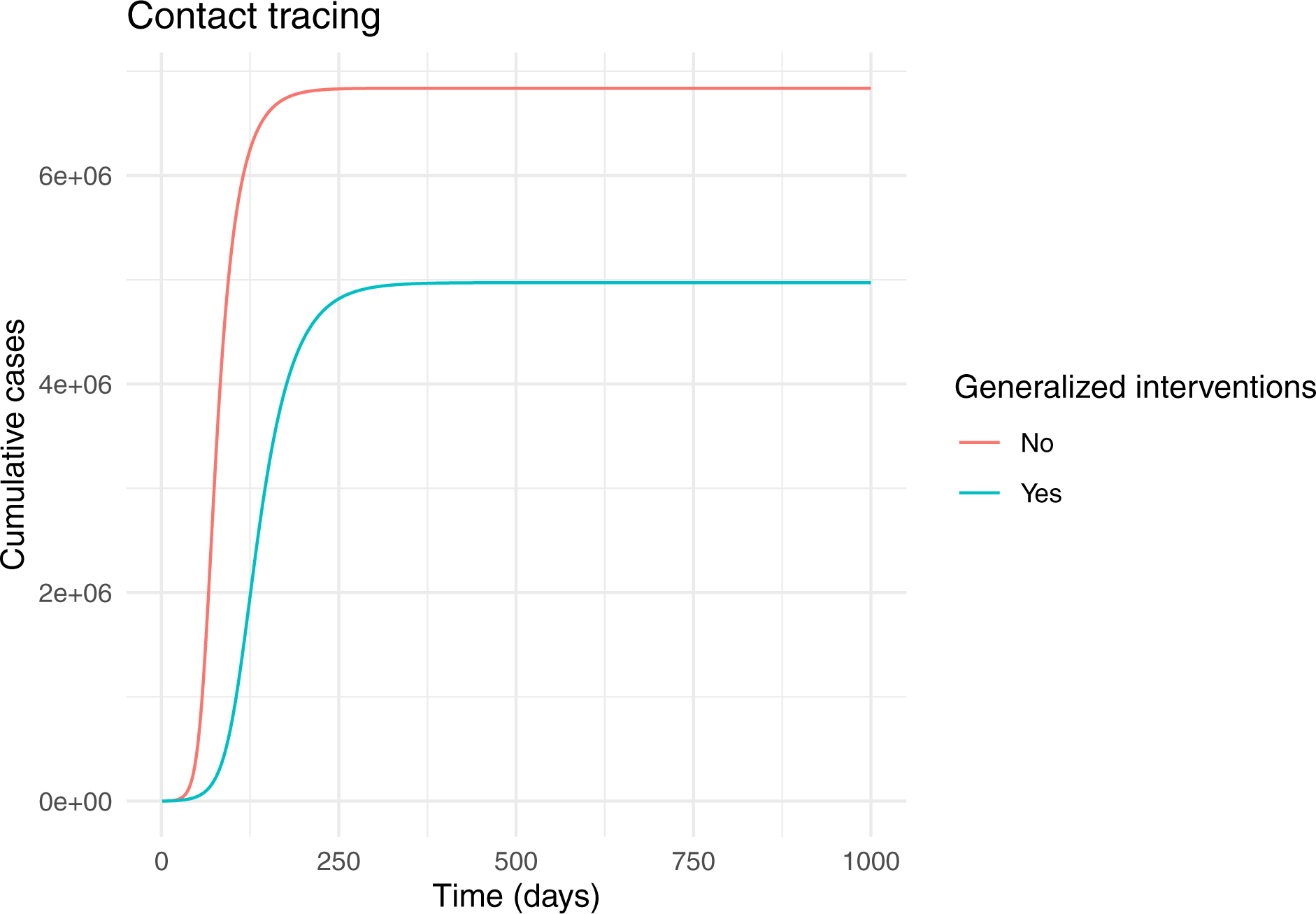

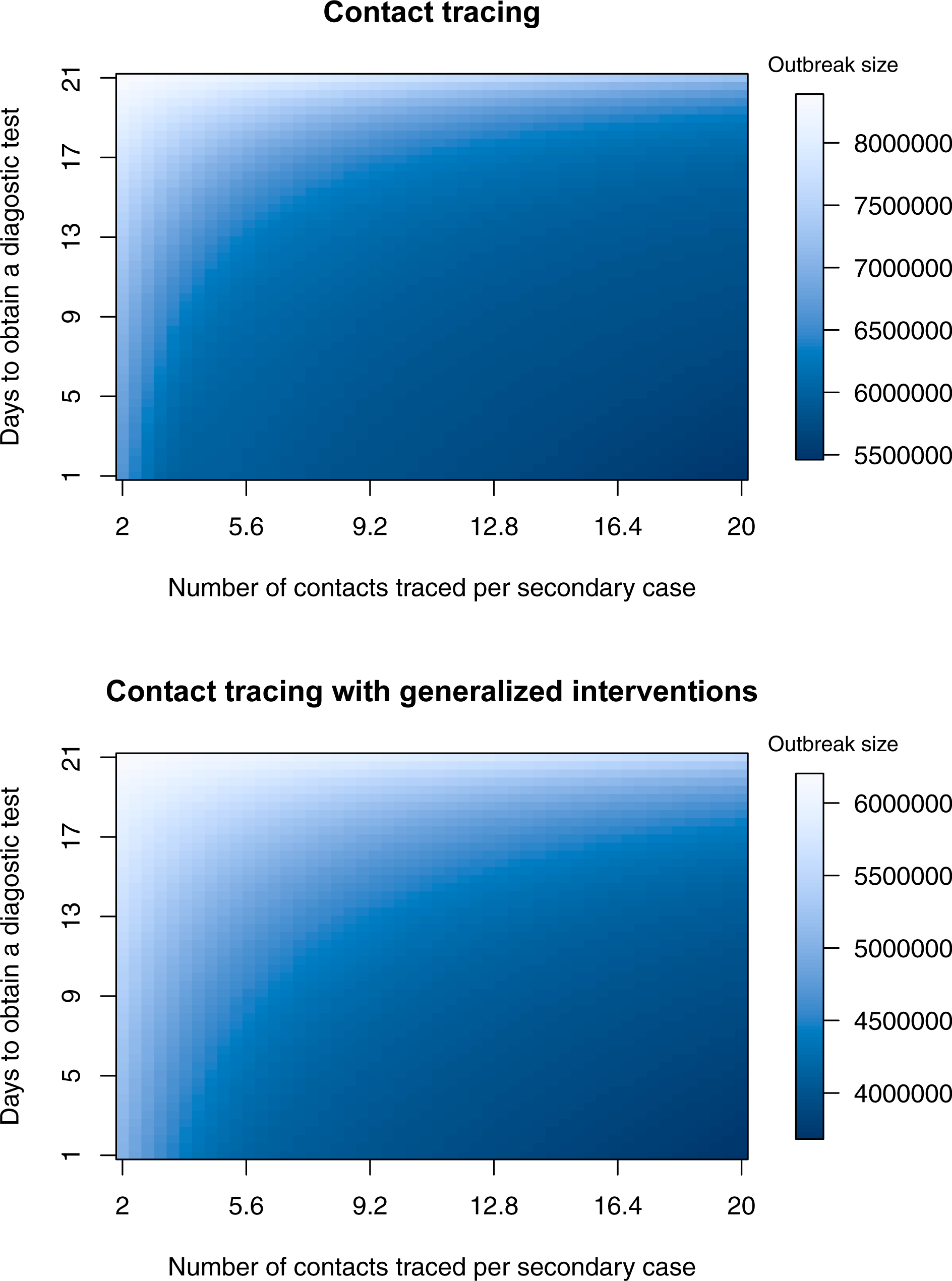

#### Contact tracing with quarantine

This scenario is the same as contact tracing except it is assumed contacts of cases are quarantined. Outcomes are qualitiatively the same and very similar in magnitude.

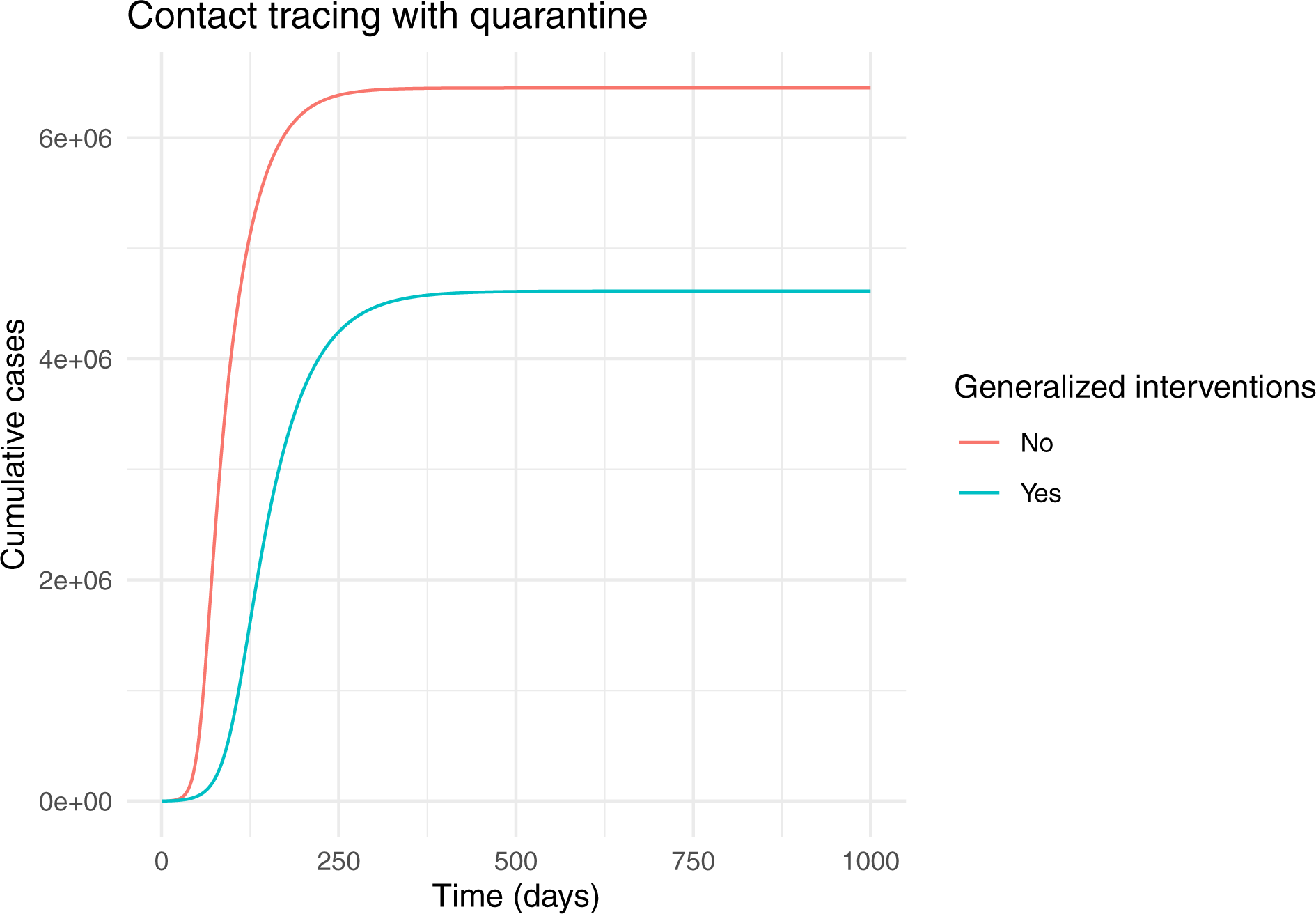

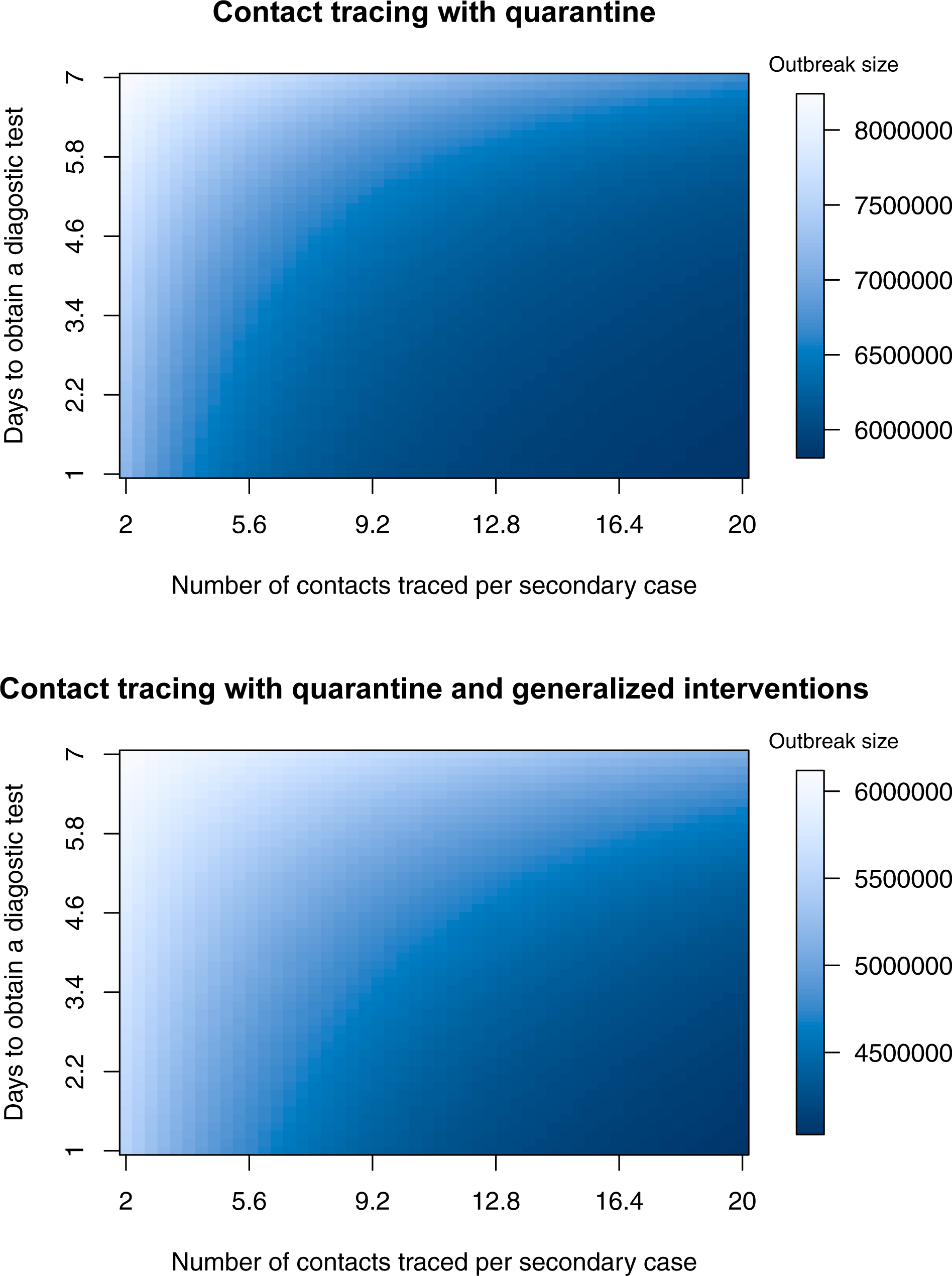

#### Certification

Here we assume that temporary certification (*ξ*) may be valid for 7 to 14 days, that it takes one to five days to obtain the results of a new test (*κ*), and that serological testing is rapid with results obtained in 14 days (*δ* = 1*/*14).

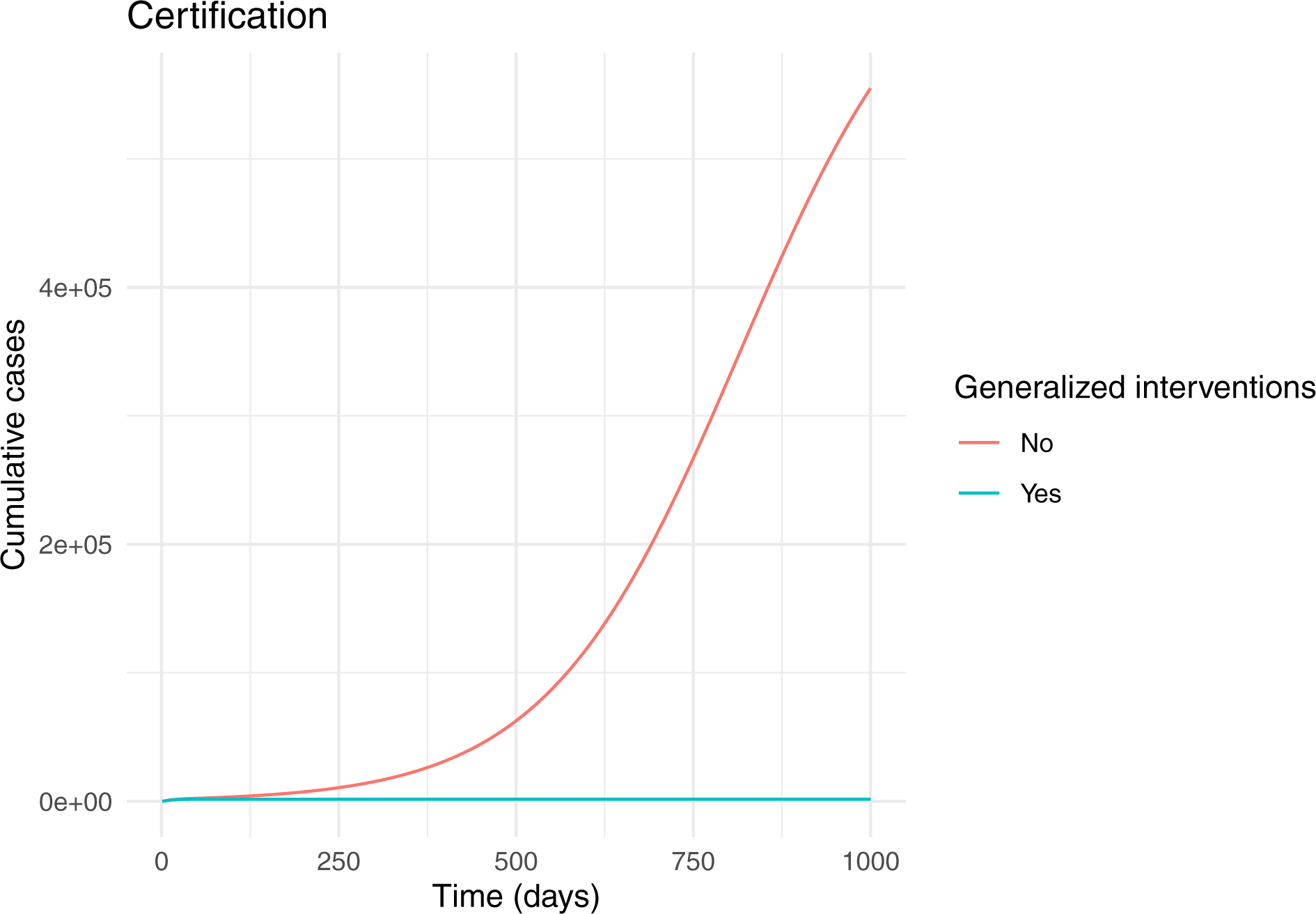

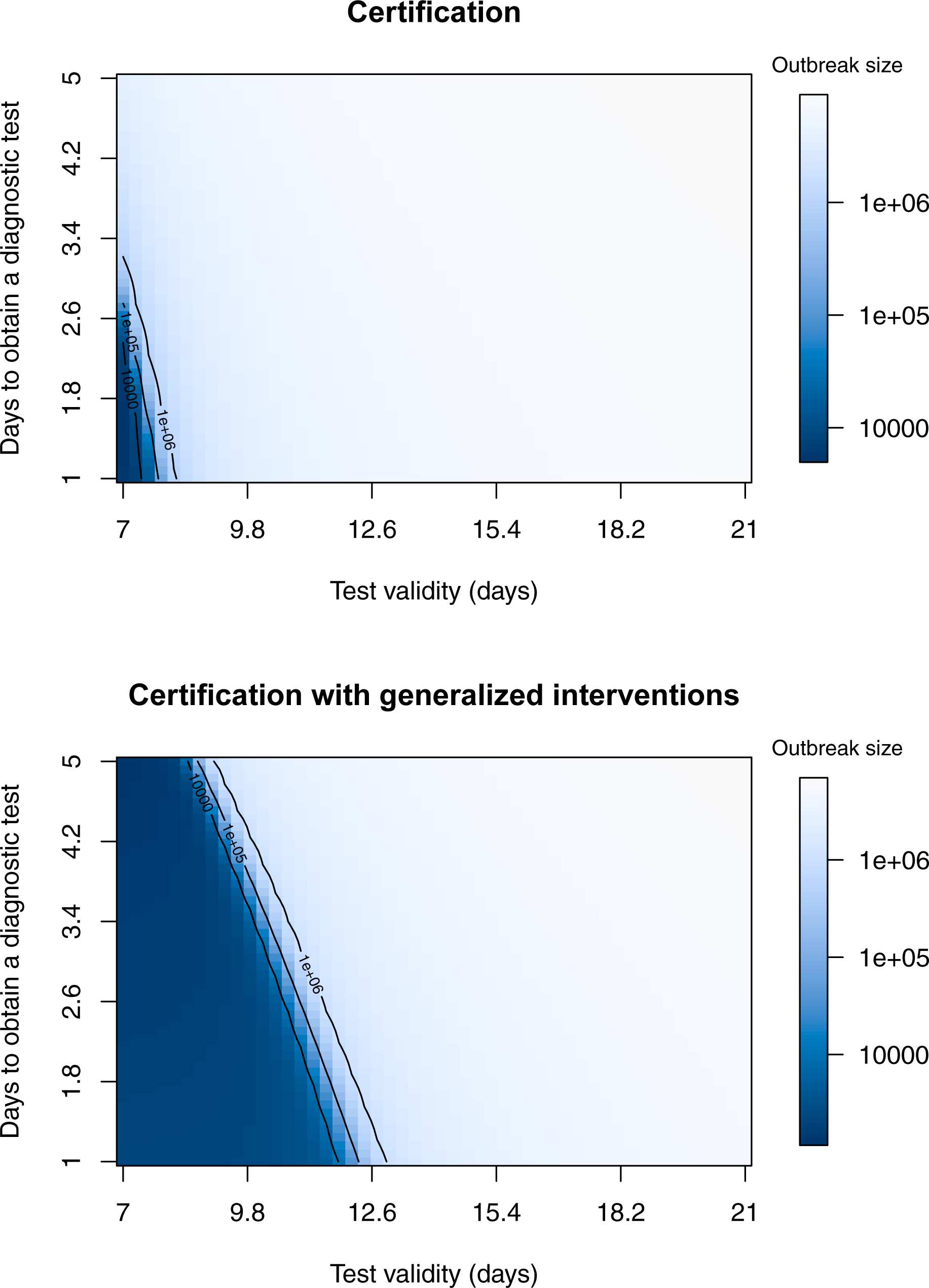

## Acknowledgements

We thank Eric Marty for assistance with figures and Paige Miller for comments on an earlier version of this paper. Andrew Tredennick assisted with troubleshooting code. Eric Marty assisted with Figures 1 and 2. Research reported here was supported by the National Institute Of General Medical Sciences of the National Institutes of Health under Award Number U01GM110744. The content is solely the responsibility of the authors and does not necessarily reflect the official views of the National Institutes of Health.

